# Regulatory architecture underlying immune dysregulation reconstructed by single-cell multi-omics in lupus nephritis

**DOI:** 10.64898/2026.05.06.26352515

**Authors:** Huanhuan Zhao, Fan Yang, Tingyu Chen, Jian Zhang, Jinsong Shi, Xiaoyang Liu, Siyi Chen, Ziyuan Ma, Shuai Liu, Xudong Fu, Na Kong, Jin Zhang, Nan Liu, Xiaomin Yu, Katalin Susztak, Xin Sheng, Zhihong Liu

## Abstract

**Objectives:** Lupus nephritis (LN) is a severe complication of systemic lupus erythematosus with heterogeneous clinical outcomes and limited therapeutic options. Although immune dysregulation is central to LN pathogenesis, the underlying cell-type-specific regulatory mechanisms and their genetic determinants remain poorly characterized.

**Methods:** We generated a single-cell multi-omics atlas of peripheral blood mononuclear cells (PBMCs) from newly diagnosed, minimally treated LN patients by integrating single-cell RNA-seq (scRNA-seq) and single-nucleus ATAC-seq (snATAC-seq) profiles. To elucidate genetically driven regulatory programs in a broaden LN population, we generated a blood expression quantitative trait loci (eQTL) atlas from 99 Chinese LN patients and performed Bayesian colocalization analysis to systematically prioritize putative causal genes for LN. Finally, we investigated how fine-mapped SNPs associated with LN phenotypic manifestations exert regulatory effects within distinct single-cell chromation contexts by leveraging peak-to-gene linkages at single-cell resolution.

**Results:** Our single-cell multi-omic dataset and orthogonal analytical approaches revealed extensive immune remodeling in LN, characterized by amplified innate immune activation and impaired adaptive immune responses, and identified transcription factors (TFs) orchestrating immune regulatory circuits. Bayesian colocalization analysis nominated 14 high-fidelity causal genes for kidney function and 23 for SLE. Integration with fine-mapped GWAS variants highlighted critical cell type convergence across autoimmune disorders and immune-mediated nephropathies, particularly within B cell subsets, where TF-driven programs delineated stage-specific differentiation networks.

**Conclusions:** Together, these analyses reconstruct the regulatory architecture underlying immune dysregulation in LN and connect genetic variation to cell-type-specific regulation, guiding genetically informed therapeutic development.

## Introduction

Lupus nephritis (LN), a severe manifestation of systemic lupus erythematosus (SLE), affects up to 70% of patients and remains a leading cause of morbidity and mortality [1]. Current treatments are limited [2], with fewer than half of patients achieving complete renal remission and 10-30% progressing to end-stage renal disease [3]. These poor outcomes largely reflect substantial heterogeneity in circulating immune profiles among LN patients [3, 4], underscoring the urgent need for refined molecular characterization to enable more effective and broadly applicable therapies.

Single-cell transcriptomic studies have revealed extensive immune heterogeneity in SLE, including aberrant type I interferon (IFN) signaling, and disrupted B cell differentiation [5–7]. However, most have primarily focused on SLE as a systemic disease [4, 5], with relatively few interrogating LN-specific immune programs [6, 7] despite its distinct pathogenic features. Concentrating on LN therefore offers an opportunity to reduce heterogeneity and enable more targeted insights into disease-specific pathogenic mechanisms.

To dissect the regulatory architecture underlying immune dysfuction in LN, it is essential to examine not only gene expression but also cis-regulatory elements (CREs) that orchestrate transcriptional programs. Single-nucleus ATAC sequencing (snATAC-seq) captures genome-wide chromatin accessibility at cellular resolution, revealing active regulatory elements and transcription factor (TF) binding motifs. Integrated with scRNA-seq, these complementary modalities yield a more complete map of immune cell identity, regulation, and dysfunction in LN.

Although analyses of gene regulation improve our understanding of disease pathogenesis, linking these regulatory circuits to genetic variation is critical for identifying causal genes relevant to the broader LN population. Prior efforts have been constrained by small sample sizes and substantial patient heterogeneity, limiting the generalizability of their findings [8]. Population-based expression quantitative trait loci (eQTL) mapping in LN patients [9, 10], provides a powerful strategy to connect genetic variants with gene expression changes, enabling the identification of high-fidelity causal genes, a strategy with strong translational value given the higher success rate of genetically supported drug targets [11]. However, blood eQTL resource for LN, particularly in the Chinese population, are currently lacking, hindering causal gene identification and targeted therapy development.

A major challenge in LN studies is disentangling disease-intrinsic molecular changes from treatment effects [3], particularly in patients with severe LN. To minimize such confounding, we profiled newly diagnosed, minimally treated LN patients using scRNA-seq and snATAC-seq of PBMCs to comprehensively map their gene regulatory programs. This analysis revealed widespread immune remodeling, characterized by heightened innate immune activity and impaired adaptive responses, driven by key TFs and cis-regulatory modules. Integrating these single-cell profiles with blood eQTL generated from 99 Chinese LN patients and fine-mapped SLE and eGFR GWAS variants prioritized 14 high-fidelity causal genes for kidney function and 23 for SLE. Fine-mapped SLE variants showed strong heritability enrichment in B cells. Further examination of TF-driven programs in B cell subsets uncovered stage-specific differentiation networks. This integrative single-cell multi-omics strategy reconstructs the regulatory architecture of immune dysregulation in LN and reveals how genetic risk variants shape cell-type-specific regulatory networks and transcriptional programs, offering valuable insights to guide genetically informed therapeutic strategies in LN.

## Methods

### Sample collection

This study was approved by the Human Subjects Committee of Jinling Hospital, Nanjing University (Approval No. 2022DZKY-060-01), and conducted according to ethical guidelines, with written informed consent obtained from all participants.

Peripheral blood was collected for scRNA-seq and snATAC-seq from 11 newly diagnosed, minimally treated female LN patients meeting the 2019 EULAR/ACR SLE criteria [12] with biopsy-confirmed LN **(Supplementary Table S1)**. Patients had no history of intravenous pulse therapy or biologic treatments within the preceding 3 months and were not receiving long-term intensive immunosuppressive therapy; hydroxychloroquine or low-to-moderate dose corticosteroids were permitted. Ten age- and sex- matched healthy donors were included as scRNA-seq controls. For eQTL analysis, whole blood from an independent cohort of 99 biopsy-confirmed LN patients was collected for bulk RNA-seq and genotyping **(Supplementary Table S2)**.

## Results

### Single-cell transcriptome profiling reveals circulating immune dysfunction in lupus nephritis

To characterize circulating immunophenotypes in LN, we performed scRNA-seq on PBMCs from 11 LN newly diagnosed, minimally treated LN patients and 10 healthy controls (HCs) **(Fig 1a and Supplementary Table S1)**. After quality control, 237,864 cells were retained and classified into 19 cell clusters using established markers [13] **(Fig 1b and** **1c**). Compared with HCs, LN patients showed reduced lymphocytes and expanded myeloid cells **(Figs 1d and** **1e** **and Supplementary Fig S1)**. These changes were also correlated with higher SLE Disease Activity Index (SLEDAI-2K) scores [14] **(Supplementary Fig S2)**.

**Figure 1.**
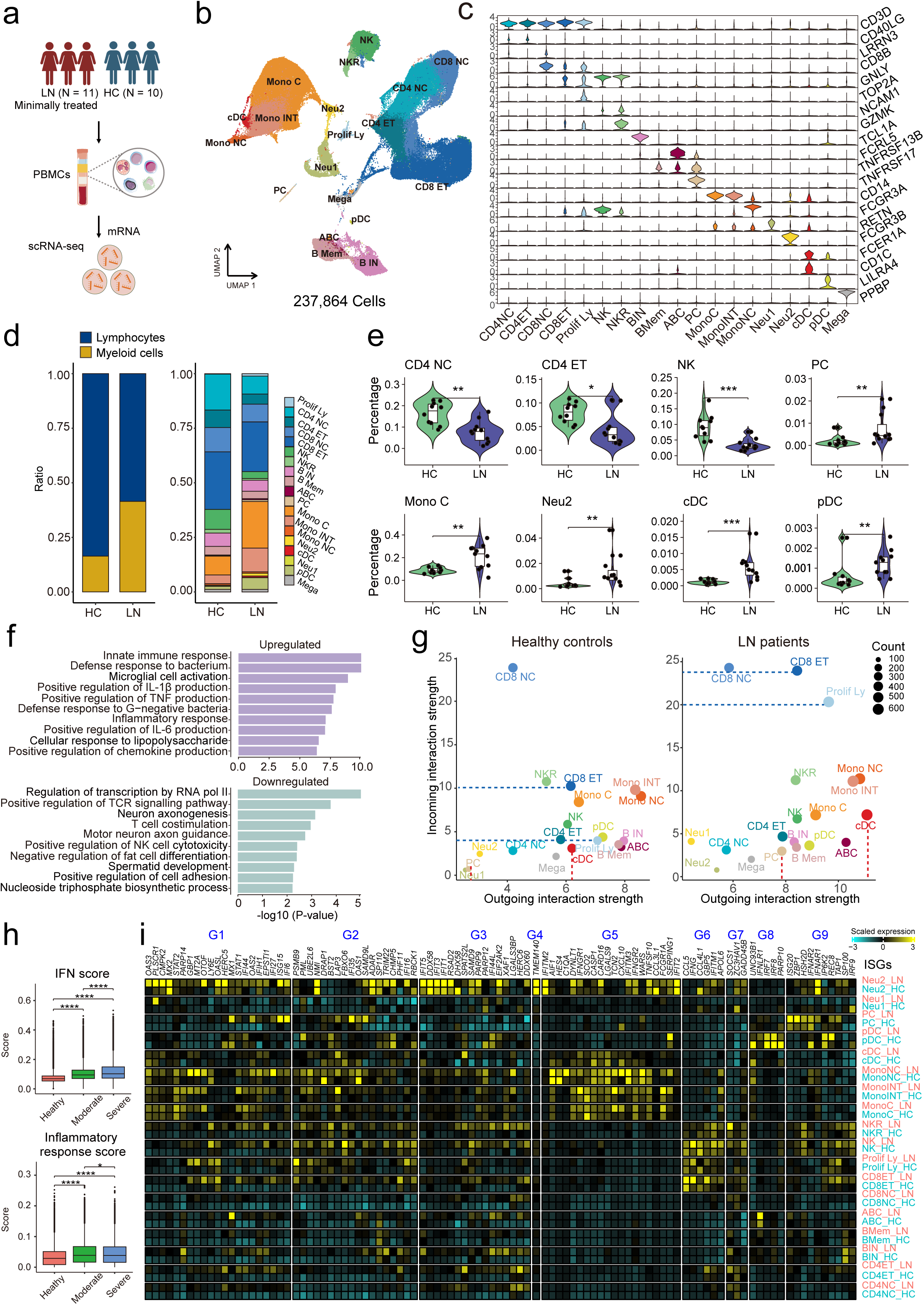
Single-cell transcriptome profiling reveals peripheral immune dysregulation in lupus nephritis. (a) Experimental scheme. scRNA-seq of PBMCs was performed on 11 newly diagnosed, minimally treated LN patients and 10 healthy controls. (b) UMAP of PBMC scRNA-seq data (n = 237,864 cells) from 11 newly diagnosed, minimally treated LN patients and 10 healthy controls. Nineteen dinstict cell clusters were identified based on estiblished cell type marker genes[13, 49]. Each data point represents a single cell, colored by cell type. CD4 NC: naïve CD4^+^ T cells, CD4 ET: effector CD4^+^ T cells, CD8 NC: naïve CD8^+^ T cells, CD8 ET: effector CD8^+^ T cells, Prolif Ly: proliferating lymphocytes, NK: natural killer cells, NKR: natural killer recruiting cells, B IN: naïve B cells, B Mem: memory B cells, ABC: age associated B cells, PC: plasma cells, Mono C: classical (CD14^++^CD16^-^) monocytes, Mono INT: intermediate (CD14^++^CD16^+^) monocytes, Mono NC: non-classical (CD14^dim^CD16^+^) monocytes, Imm_Neu: immature neutrophils, Neu: neutrophils, cDC: conventional dendritic cells, pDC: plasmacytoid dendritic cells, Mega: megakaryocytes. (c) Violin plots showing the expression levels of representative cell type markes. *Y*-axis shows log-scale normalized read count. (d) Bar plots showing relative cell abundances in healthy controls and LN patients. Colors indicate cell lineages (*right*) and individual cell types (*left*). (e) Violin plots showing cell fractions in healthy controls (HC, n = 10) and LN patients (n = 11). P-values were calculated using the Wilcoxon test comparing group means between LN and HC. Significance levels are indicated as *p < 0.05, **p < 0.01, ***p < 0.001, and ****p < 0.0001. CD4 NC: naïve CD4^+^ T cells, CD4 ET: effector CD4^+^ T cells, NK: natural killer cells, PC: plasma cells, Mono C: classical (CD14^++^CD16^-^) monocytes, Neu: neutrophils, cDC: conventional dendritic cells, pDC: plasmacytoid dendritic cells. The center lines show the medians; the box limits indicate the 25^th^ and 75th percentiles; the whiskers extend to the 5th and 95th percentiles; the outliers are represented by the dots. (f) Functional annotation (gene ontology) of the 1,939 upregulated genes *(top)* and 1,285 downregulated genes *(bottom)* in LN patients. (g) Scatter plot showing outgoing interaction strength (*X*-axis) and incoming interaction strength (*Y*-axis) for each immune cell type in HC (*left*) and LN (*right*). Each dot represents a cell population, colored by cell identity, with dot size reflecting the number of inferred interactions. Dashed blue horizontal and red vertical lines highlight cell types with high incoming and outgoing interaction strengths, respectively. (h) Interferon (IFN) scores (*top*) and inflammatory response (IR) scores (*bottom*) of PBMCs in HC, moderate LN (AI ≤ 7), and severe LN (AI > 7). The center lines show the medians; the box limits indicate the 25th and 75th percentiles; the whiskers extend to the 5th and 95th percentiles; the outliers are represented by the dots. P-value was calculated using two-sided t-test with Bonferroni-adjusted P-values. Significance levels are indicated as *p < 0.05, **p < 0.01, ***p < 0.001, and ****p < 0.0001. (i) Heatmap showing row-scaled expression of ISGs (n = 100 unique genes) across 18 cell clusters (megakaryocytes excluded) in healthy controls and LN patients. The color scheme is based on Z-scores, calculated from normalized gene expression levels of the 18 cell types. Z-scores show the relatively expression levels compared to other clusters. Gene ordering and cluster groupings (G1-G9) are identical to those reported in the recent PBMC scRNA-seq study by Nehar-Belaid et al[5].

Pseudobulk differential gene expression (DEG) analysis identified 1,939 upregulated and 1,285 downregulated genes in LN PBMCs **(Supplementary Fig S3 and Supplementary Table S3)**. Upregulated genes were enriched for innate immune response, whereas downregulated genes were enriched for T cell receptor signaling pathway, costimulation, and NK cell-mediated cytotoxicity **(Fig 1f and Supplementary Table S4 and S5)**. Downregulated genes were predominantly expressed in lymphocytes and linked to lymphocyte proliferation pathways, whereas upregulated genes were enriched in myeloid cells and proliferating lymphocytes. Intriguingly, the expression patterns of the top DEGs displayed strong cell-type specificity **(Supplementary Fig S4)**. Cell-cell interaction analysis revealed increased incoming signals to effector CD8^+^ T cells and proliferating lymphocytes, alongside enhanced outgoing signals from conventional dendritic cells and plasma cells (PCs) **(Fig 1g and Supplementary Fig S5)**.

Stratifying LN patients by kidney activity index (AI) [15] into moderate (AI ≤ 7) and severe (AI > 7), we assessed enrichment scores of SLE-related interferon-stimulated genes (ISGs) [5] and inflammatory genes [6] **(Supplementary Table S6 and Supplementary Note)**. IFN and inflammatory response scores were higher in LN than HCs **(Fig 1h)**, with IFN scores increasing with disease severity **(Fig 1h and Supplementary Fig S6a)**. Monocytes and neutrophils showed the strongest increase in IFN scores, while PCs and proliferating lymphocytes exhibited the greatest reduction in inflammatory response scores **(Supplementary Figs S6b and S6c)**. ISG scores also rose in naïve B cells (B IN), memory B cells (B Mem), age-associated B cells (ABCs), and effector CD8^+^ T cells, despite stable cell proportions **(Supplementary Fig S1b)**.

Next, we examined single-cell expression of ISGs and pro-inflammatory genes in LN patients and HCs **(Fig 1i and Supplementary Fig S7)**. Using previously defined ISG clusters (G1-G9) from SLE PBMC scRNA-seq [5], we observed similar cell-type- and disease-specific expression patterns, including upregulation of *STAT2*, *IFIT5*, *IFI44L*, and *XAF1* in B IN, and *MX1* and *IFNLR1* in ABCs in LN patients **(Fig 1i)**. The strongest ISG upregulation occured in G1, particularly in neutrophil subpopulation 2 (Neu_2) and monocytes.

### Single nuclei chromatin accessibility profiling identifies lineage-specific regulatory signatures in LN PBMCs

In parallel, to investigate epigenomic regulatory programs underlying transcriptomic changes, we performed snATAC-seq on PBMCs from the same 11 LN patients **(Fig 2a and Supplementary Table S1)**. After quality control, 45,107 high-quality cells from 9 LN patients were retained. Based on promoter accessibility of known markers [13], we identified 242,637 accessible chromatin peaks across 15 immune cell types, enriched in promoter and distal regulatory regions **(Fig 2b and Supplementary Fig S8)**. Chromatin accessibility of key markers showed clear cell-type specificity **(Fig 2c)**, consistent with scRNA-seq expression patterns **(Fig 1c)**. Gene activity scores for the top 3,000 variable genes across 14 matched cell types strongly concorded with scRNA-seq **(Fig 2d)**. Cell composition was also highly consistent between datasets, with reduced lymphocytes and increased myeloid cells correlating with higher SLEDAI-2K scores **(Fig 2e)**.

**Figure 2.**
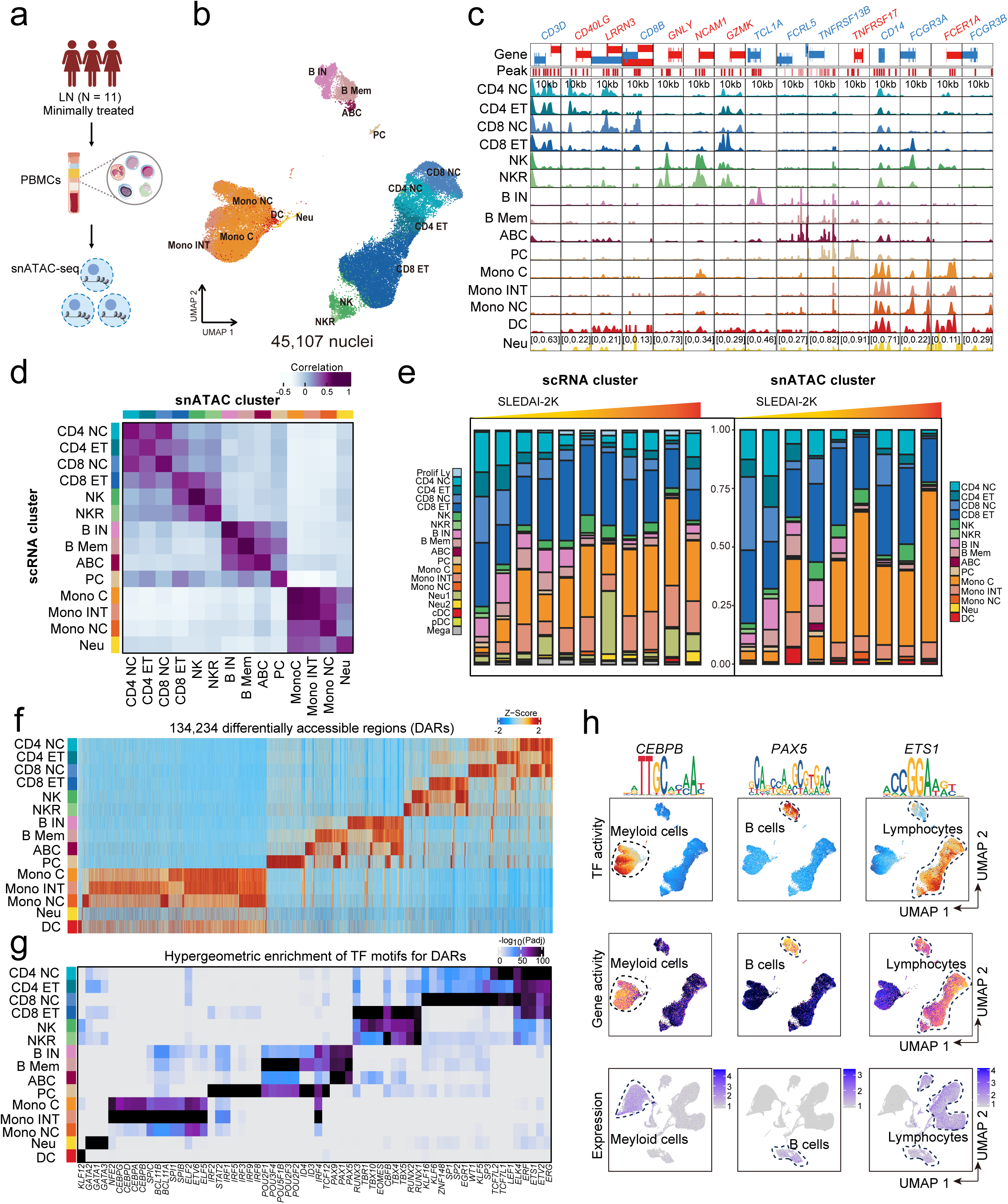
Single-nucleus chromatin accessibility reveals lineage-specific regulatory programs in LN PBMCs. (a) Experimental scheme. snATAC-seq of PBMCs was performed on the same cohort of 11 newly diagnosed, minimally treated LN patients as used for scRNA-seq. (b) UMAP of PBMC snATAC-seq data (n = 45,107 nuclei) from the same 9 newly diagnosed, minimally treated LN patients, with 2 low-quality samples excluded. Fifteen dinstict cell clusters were identified based on estiblished cell type marker genes[13, 49]. Each data point represents a single cell, colored by cell type. CD4 NC: naïve CD4^+^ T cells, CD4 ET: effector CD4^+^ T cells, CD8 NC: naïve CD8^+^ T cells, CD8 ET: effector CD8^+^ T cells, NK: natural killer cells, NKR: natural killer recruiting cells, B IN: naïve B cells, B Mem: memory B cells, ABC: age associated B cells, PC: plasma cells, Mono C: classical (CD14^++^CD16^-^) monocytes, Mono INT: intermediate (CD14^++^CD16^+^) monocytes, Mono NC: non-classical (CD14^dim^CD16^+^) monocytes, DC: dendritic cells, Neu: neutrophils. (c) Genome browser plots showing aggregate read densities of representative cell type marker genes: *CD40LG^+^LRRN3^+^* (naïve CD4^+^ T cell); *CD40LG^+^GZMK^+^* (effector CD4^+^ T cell); *CD8B^+^LRNN3*^+^ (naïve CD8^+^ T cell); *CD8B^+^GZMK^+^* (effector CD8^+^ T cell); *NCAM1* (natural killer cell); *NCAM1^+^GZMK^+^*(natural killer recruiting cell); *TCL1A* (naïve B cell); *TNFRSF13B* (memory B cell); *FCRL5* (age associated B cell); *TNFRSF17* (plasma cell); *CD14*^++^ (classical monocyte); *CD14^+^FCGR3A^+^* (intermediate monocyte); *FCGR3A^++^* (non-classical monocyte); *FCER1A* (dendritic cell); and *FCGR3B* (neutrophil). (d) Heatmap showing Pearson’s correlation between gene activity scores from snATAC-seq and gene expression values from scRNA-seq in LN patients. The color scheme is based on Pearson’s correlation coefficients. The color keys along the left column and top row denote cell clusters identified by scRNA-seq and snATAC-seq, respectively. (e) Bar plots showing relative cell abundances across LN patients in scRNA-seq (*left*) and snATAC-seq (*right*) datasets. Patients are ordered by increasing SLEDAI-2K scores[14]. Colors denote individual cell types. (f) Heatmap showing Z-score normalized accessibility of 134,234 differentially accessible regions (DARs) across 15 immune cell types. The color scale indicates Z-scores, with red representing high accessibility and blue representing low accessibility. (g) Heatmap showing hypergeometric enrichment of TF motifs within DARs across 15 immune cell types. The color scale represents normalized enrichment as -log_10_(P_adj_), with darker shades indicating stronger and more significant enrichment. (h) UMAP showing TF activity scores (*top*) and gene activity scores (*middle*) estimated from snATAC-seq, as well as gene expression levels from scRNA-seq (*bottom*), for *CEBPB* (*left*), *PAX5* (*middle*), and *ETS1* (*right*).

Next, we identified 134,234 differentially accessible regions (DARs) across cell clusters **(Fig 2f)**. Motif enrichment analysis identified lineage-determining TFs in cell-type-specific DARs **(Supplementary Table S7 and Fig 2g)**. Key TFs showed concordant motif activity, chromatin accessibility, and expression, such as *CEBPB* in myeloid cells, *PAX5* in B cells, and *ETS1* in lymphocytes **(Fig 2h)**.

### Single-cell integration of snATAC-seq and scRNA-seq maps CRE-gene regulatory modules in LN

Next, we integrated snATAC-seq and scRNA-seq across shared cell types using canonical correlation analysis (CCA) to align chromatin accessibility with gene expression, and inferred CREs by correlating chromatin accessibility peaks with local gene expression (‘peak-to-gene’ linkages) [16] **(Fig 3a)**. Linkages were identified using the entire PBMC dataset and within individual immune cell lineages. We identified 91,479 peak-to-gene linkages with scores > 0.5 **(Supplementary Note)**, of which only 45,281 were detected when using the entire PBMC dataset alone **(Supplementary Fig S9a)**. Over 56.97% were B cell-specific, whereas only a small subset showed specificity to myeloid cells. A greater number of identified linkages were supported by enhancer-gene pair predictions from a large activity-by-contact model dataset [17, 18] **(Supplementary Fig S9b)**, had higher GC content, and were farther from the transcription start sites (TSSs) of their associated genes than unlinked peaks **(Supplementary Figs S9c and S9d)**. Of 242,637 accessible peaks, 37.70% were linked to genes, and 39.10% to the nearest gene, consistent with previous experimental estimates [18] **(Supplementary Fig S9e)**. We next identified 1,178 highly regulated genes (HRGs) by ranking genes based on the number of linked CREs and retaining those with ≥ 20 peak-to-gene linkages **(Fig 3b and Supplementary Table S8)**, whose aggregate CRE accessibility strongly correlated with expression **(Supplementary Fig S9f)**. HRGs were enriched for super-enhancer-associated genes [19] **(Fig 3c and Supplementary Table S9)**, B cell lineage factors (*BACH2*, *PRDM1*) [20], and type I interferon signaling components (*IFNAR1*, *IRF5*) [21] **(Fig 3b)**. In LN PBMCs, 90 HRGs were downregulated and 632 upregulated **(Fig 3d and Supplementary Table S3)**. Downregulated HRGs were primarily involved in pathways related to TCR signaling **(Fig 3d and Supplementary Table S10)**, whereas upregulated HRGs were enriched for innate immune activation and inflammatory pathways **(Fig 3d and Supplementary Table S11)**. *K*-nearest neighbor clustering of pseudobulk CREs for HRGs revealed a spectrum from highly cell-type-specific to broadly shared CREs. For example, while *TCF7*-linked CREs were widely shared, *IRF8*- and *PRDM1*-linked CREs formed distinct cell-type specific modules **(Fig 3e)**.

**Figure 3.**
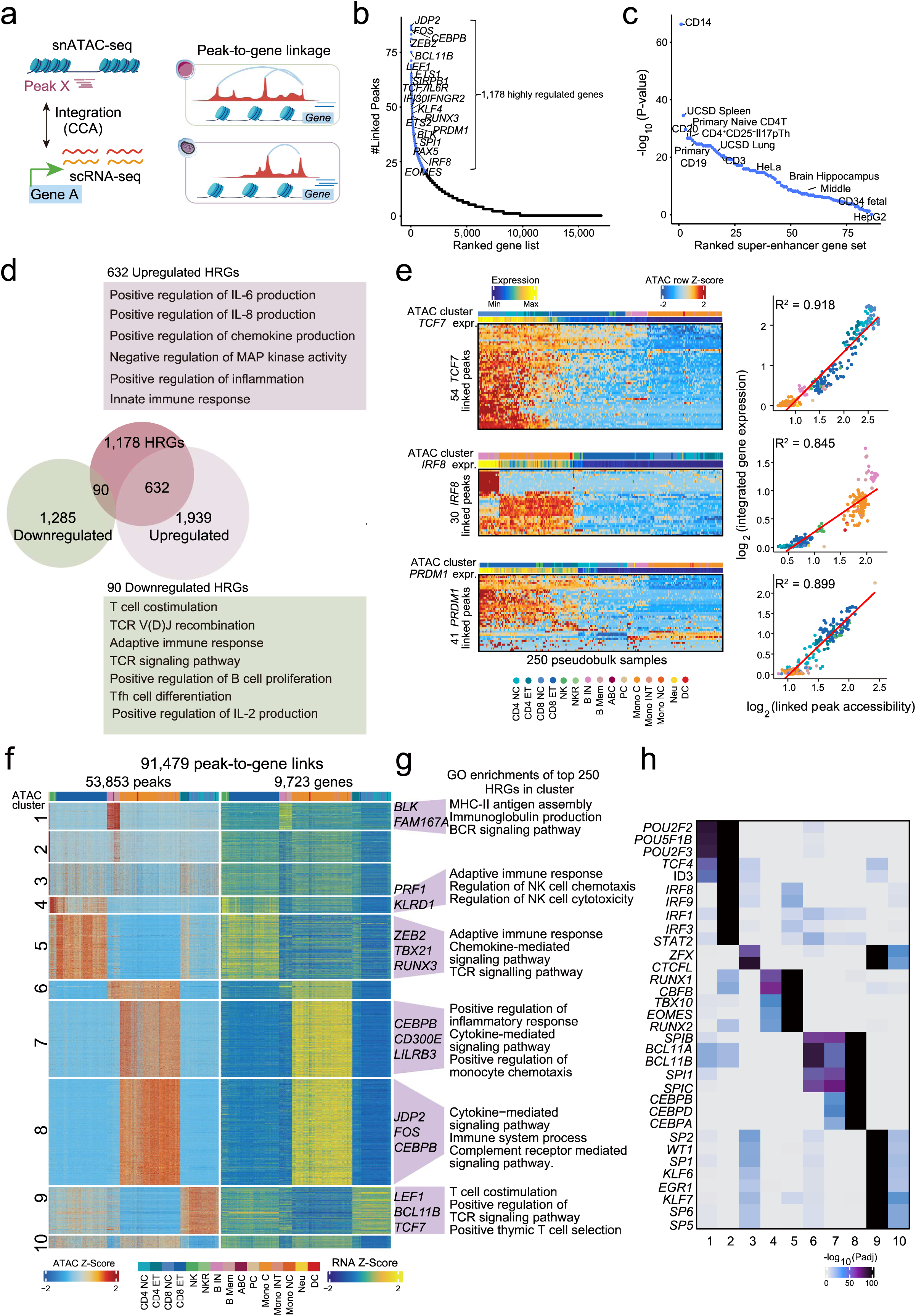
Single-cell integration of chromatin accessibility and gene expression uncovers CRE-gene regulatory modules in LN. (a) Experimental scheme. Chromatin accessibility was aligned with gene expression across shared cell types from snATAC-seq and scRNA-seq using canonical correlation analysis (CCA)[16], a method for multi-modal data integration. Cis-regulatory elements (CREs) were then inferred by correlating chromatin accessibility peaks with local gene expression, generating ‘peak-to-gene’ linkages. (b) Genes ranked by the number of peak-to-gene linkages per gene. A total of 1,178□HRGs with >20□ linkages were identified. Each dot represents a single gene. (c) Hypergeometric enrichment of super-enhancer-linked genes among HRGs across multiple cell and tissue types. Each point represents a cell type or tissue. (d) Overlap of 1,178 HRGs with differentially expressed genes from LN patients and healthy controls (*left*). Functional annotation (gene ontology) of 90 downregulated HRGs (*top right*) and 632 upregulated HRGs (*bottom right*). (e) Heatmap showing chromatin accessibility at *TCF7* (*top*), *IRF8* (*middle*), and *PRDM1* (*bottom*) linked peaks across 250□pseudobulked snATAC-seq samples (*left*). Cell type labels are shown above the heatmap, with corresponding gene expression levels displayed below. Scatter plots (*right*) showing the relationship between linked peak accessibility and gene expression for each pseudobulked samples, with the red line indicating the line of best fit. (f) Heatmaps of chromatin accessibility (*left*) and gene expression (*right*) for 91,479 peak-to-gene linkages, clustered by *k*-means (*k*□=□10). Representative HRGs for selected clusters are shown to the right of the gene expression heatmap. Rows indicate scaled accessibility (Z-score, *left*) and gene expression (Z-score, *right*). Each column represents a pseudobulk sample, with the top color bar denoting cluster identity. (g) Gene ontology enrichment for the top 250□genes ranked by peak-to-gene linkage number in selected *k*-means clusters. (h) Heatmap showing hypergeometric enrichment of TF motifs within peaks from each *k*-means cluster (k=10), represented as -log_10_(adjusted P-value).

To explore regulatory heterogeneity in LN, we performed *k*-means clustering (k = 10) to CRE accessibility from 91,479 peak-to-gene linkages, identifying cell-type-specific regulatory modules **(Fig 3f)**. For instance, cluster 1 was B cell-specific, with linked genes enriched for immunoglobulin production **(Fig 3g, Supplementary Tables S12 and S13)**. Clusters 7 and 8 were myeloid-specific, enriched for genes regulating monocyte chemotaxis and cytokine-mediated signaling pathways. Motif enrichment analysis of clustered CREs highlighted *POU2F2*, a regulator of B cell proliferation and differentiation [22], enriched in clusters 1 and 2 **(Fig 3h)**.

### Key transcriptional regulators orchestrating immunological circuits rewiring in LN

To identify TFs driving immune rewiring in LN, we performed motif enrichment on CREs and correlated TF expression with the overall accessibility of their associated target genes [23] **(Fig 4a)**. This revealed 20 TFs associated with downregulation of 69 HRGs and 35 TFs associated with upregulation of 621 HRGs **(Figs 4b and** **4c****, Supplementary Tables S14 and S15)**. *LEF1* and *TCF7* were central activators of downregulated HRGs, with *KLF10* and *EGR1* as repressors **(Fig 4b and Supplementary Fig S10a)**. Upregulated HRGs were controlled by activators governing myeloid cell differentiation [24] (*CEBPA/B*, *SPI1*) and repressors regulating lymphocyte development and differentiation [25] (*BCL11B*, *BACH2*) **(Fig 4c and Supplementary Fig S10b)**. Notably, *SPI1* and *BCL11B* displayed enriched motif activity in myeloid cells and B cells, but only *SPI1* showed concordantly high gene activity and expression within the same cell clusters [26] **(Fig 4d)**. As expected, key activators of downregulated HRGs (*LEF1*, *TCF7*) and repressors of upregulated HRGs (*BCL11B*, *BACH2*) were higher in HCs, whereas repressors of downregulated HRGs (*KLF10*, *EGR1*) and activators of upregulated HRGs (*SPI1*, *CEBPA*) were elevated in LN **(Fig 4e)**. We also examined regulators of SLE-associated ISGs [5] **(Supplementary Table S16)** and found that *SPI1*, *CEBPA*/*B*, *BCL11B*, and *SPIB* also controlled ISG expression **(Fig 4c,Supplementary Fig S11)**.

**Figure 4.**
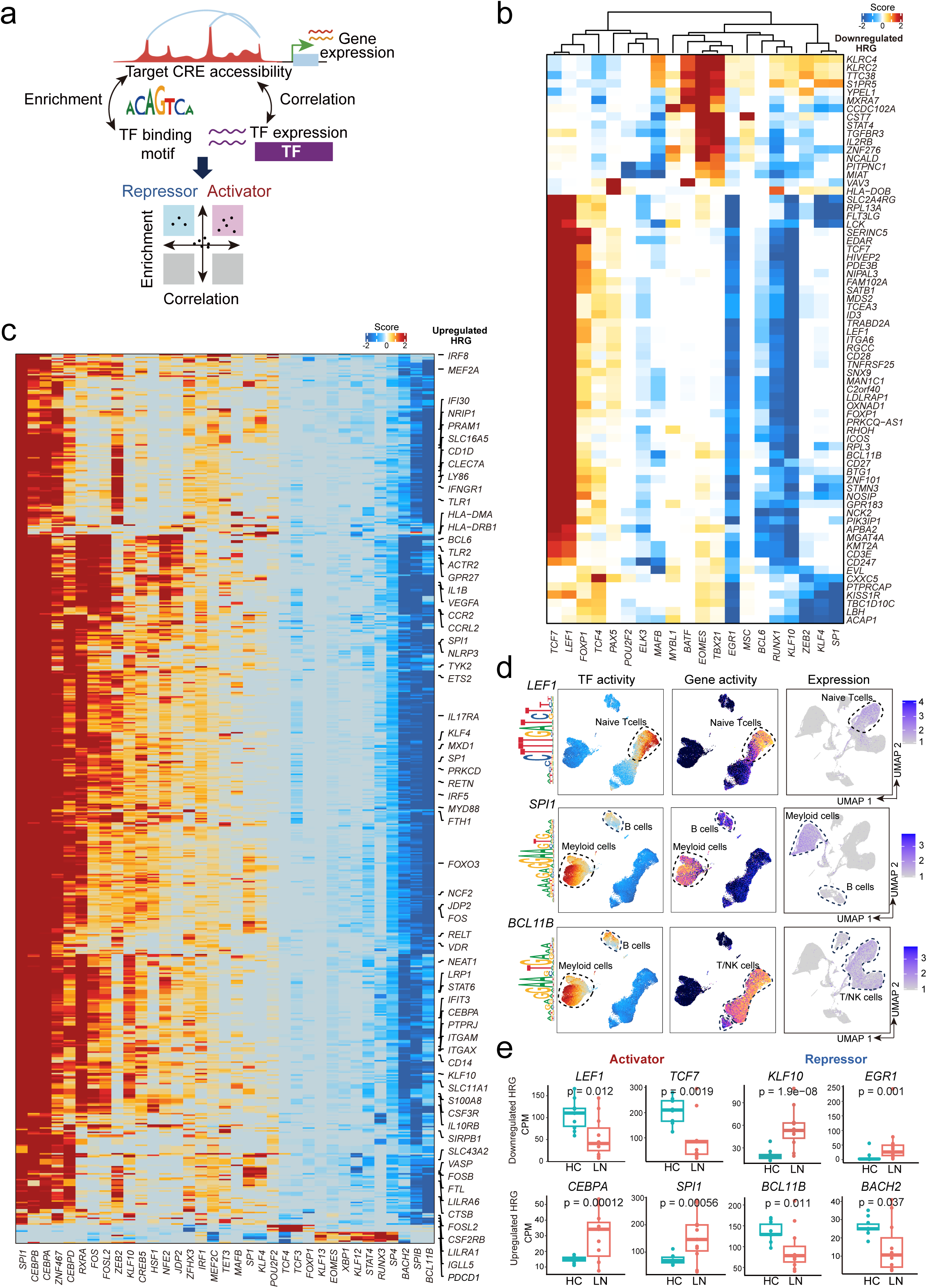
Identification of transcriptional regulators driving immunogical circuit rewiring in lupus nephritis. (a) Experimental scheme. Potential transcriptional activators and repressors were identified by performing motif enrichment analysis on CREs and assessing the correlation between TF expression and the overall accessibility of their associated target genes. (b) Heatmap showing regulation scores for TFs associated with downregulated HRGs. Positive and negative regulatory effects are shown in red and blue, respectively. Rows represent 69 target genes and columns represent 20 TFs. (c) Heatmap showing regulation scores for TFs associated with upregulated HRGs. Positive and negative regulatory effects are shown in red and blue, respectively. Rows represent 621 target genes and columns represent 35 TFs. (d) UMAP showing TF activity scores (*left*) and gene activity scores (*middle*) estimated from snATAC-seq, as well as gene expression levels from scRNA-seq (*right*), for *LEF1* (*top*), *SPI1* (*middle*), and *BCL11B* (*bottom*). (e) Representative examples of differentally expressed HRGs between 10 healthy controls and 11 LN patients. Pseudobulk expression (counts per million, CPM) was calculated by aggregating raw scRNA-seq counts across cells for each individual. Statistical significance was assessed using the Wilcoxon rank-sum test, with two-sided P-values adjusted by the Benjamini-Hochberg method. The center lines show the medians; the box limits indicate the 25th and 75th percentiles; the whiskers extend to the 5th and 95th percentiles; the outliers are represented by the dots.

### Mapping genetic regulation of TF-gene networks across the LN blood transcriptome

To define population-level TF-gene regulattion, we generated a whole-blood eQTL atlas from 99 Chinese LN patients **(Fig 5a and Supplementary Table S2)** using the cell fraction eQTL(cf) model to account for cell composition effects, which we previously showed markedly improves eQTL discovery power [9] **(Supplementary Fig S12a-c and Supplementary Note)**. We identified 1,205 eGenes (genes associated with at least one SNP) and 107,701 significant SNP-gene pairs [27] **(Supplementary Table S17)**. Our results showed strong replication of previously identified SNP-gene associations in whole blood from the GTEx [28] with π_1_ = 0.986 (where π_1_ represents the fraction of true positives [29]), consistent with our own and others’ previous identification of eQTLs from the same tissue [9, 10, 30]. The number of identified eGenes aligns with previous eQTL studies across 49 human tissues from the GTEx Consortium [28] **(Supplementary Fig S12d)**. For instance, the genetic variants rs12928014 and rs7313235 showed robust effects on *PRKCB* and *CLEC12A* expression, respectively **(Fig 5b)**. 113 eGenes overlapped with HRGs from integrated scRNA-seq and snATAC-seq data **(Fig 5c)**, primarily expressed in myeloid cells **(Supplementary Fig S13)**. eQTL(cf) SNP-induced motif disruptions highlighted *SPI1, CEBPA,* and *LEF1* **(Fig 5d and Supplementary Table S18)**. Approximately 10.9% of CREs overlapped eQTL(cf) SNPs, and 14.8% were linked to the same eGenes, with the strongest enrichment in monocyte- and B cell-specific CREs **(Fig 5e and Supplementary Fig S14)**.

**Figure 5.**
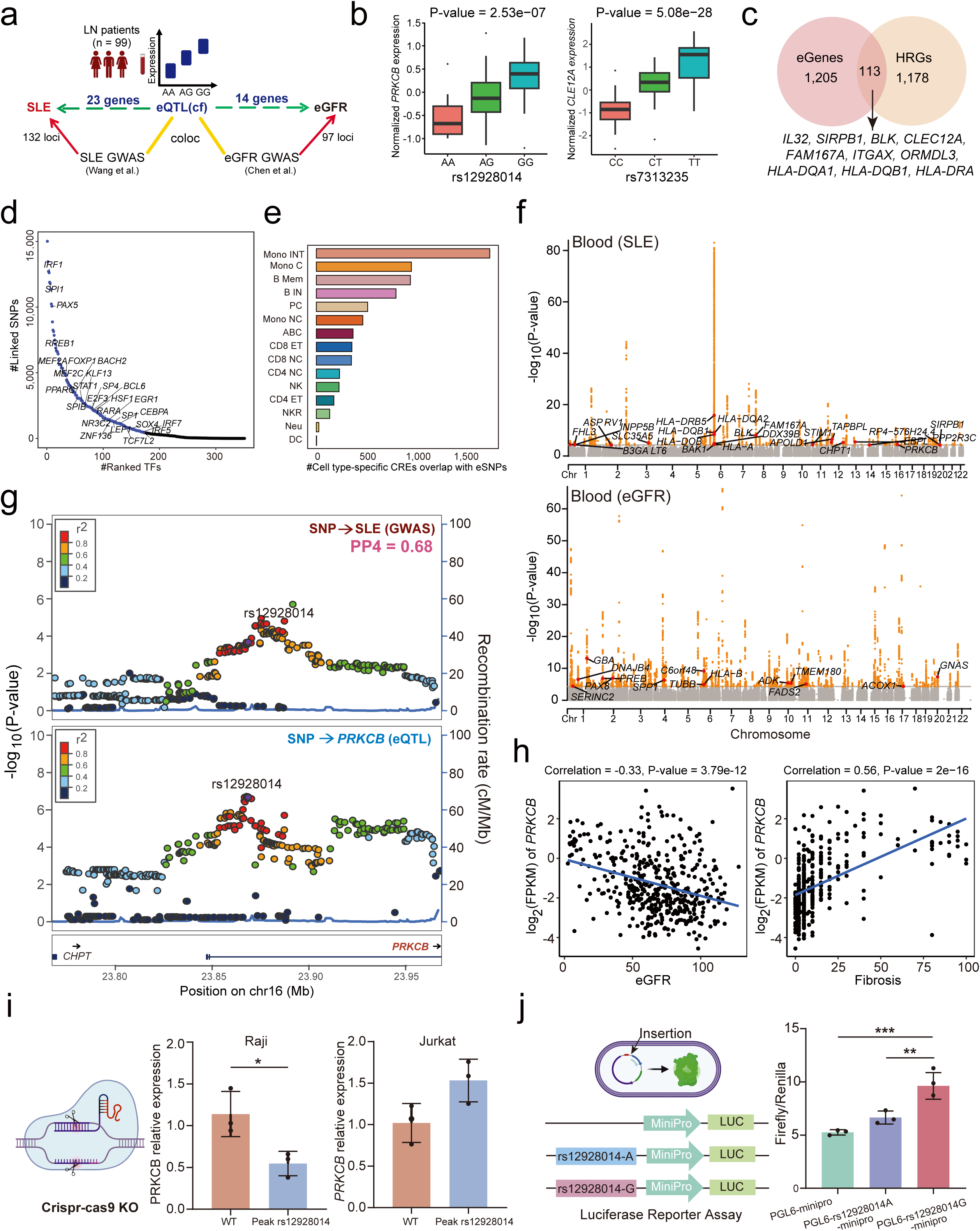
eQTL mapping and colocalization prioritize candidate causal genes in lupus nephritis. (a) Experimental scheme. We generated a comprehensive whole blood eQTL atlas from 99 Chinese LN patients by mapping genotype effects on gene expression using the cell fraction eQTL(cf) model. To prioritize candidate genes contributing to LN susceptibility, we performed Bayesian colocalization analysis by integrating Asian SLE and eGFR GWAS summary statistics with whole blood eQTL(cf) data from LN patients. This approach identified loci where causal genetic variants were shared between gene expression changes and LN susceptibility. (b) Association between the SNP genotypes and gene expression in blood samples (n = 99). The *y* axis shows the *PRKCB*- (*left*) and *CLEC12A-* (*right*) normalized expression in blood; the *x* axis indicates genotypes at the rs12928014 (*left*) and rs7313235 (*right*) loci. The center lines show the medians; the box limits indicate the 25th and 75th percentiles; the whiskers extend to the 5th and 95th percentiles; the outliers are represented by the dots. The two-sided P-value was calculated using the linear regression eQTL(cf) model. (c) The number of eGenes influenced by at least one genetic variant, as identified by eQTL analysis. HRGs were defined as genes with more than 20 peak-to-gene linkages. Among them, 113 eGenes overlapped with HRGs, suggesting that their regulation could be influence by underlying genetic variants. (d) Putative TF-binding motifs disrupted by eSNPs, ranked by the number of linked eSNPs. A total of 1,178□HRGs with more than 20□ linkages were identified. Each dot represents a TF-binding motif disrupted by eSNPs. Blue dots denote the top 50% of motifs, each disrupted by at least 425 linked eSNPs. (e) Bar plots showing the overlap between eSNPs with cell type-specific CREs across immune cell types. Colors denote individual cell types. (f) Miami plot showing genes prioritized by colocalization[33] of SLE[31] (*top*) and eGFR[32] (*bottom*) GWAS signals with blood eQTL(cf)s in LN patients. The red dots define prioritized protein-coding genes. The *x* axis represents the chromosome. The *y* axis represents -log_10_(P). The two-sided P-value was obtained from the GWAS study[31, 32]. (g) LocusZoom plots of SLE GWAS variants (*top*) and blood *PRKCB* eQTL(cf)s (*bottom*). The *x* axis shows the ±100 kb genomic region around rs12928014. The *y* axis represents the significance -log_10_(two-sided P-value) of association tests (by linear regression). Each data point represents a variant, with color denoting the r^2^ (the degree of LD). n = 12,653 individuals for SLE GWAS; n = 99 individuals for LN blood eQTL(cf)s. (h) Pearson’s correlation of *PRKCB* expression with eGFR (*left*) and fibrosis (*right*) across 433 micro-dissected human kidney tubular samples. Two-sided P-value was calculated by t-test. (i) CRISPR-Cas9 knockout of the rs12928014-overlaping region alters *PRKCB* expression. Relative *PRKCB* transcript levels in Raji and Jurkat cells with wild-type (WT) or CRISPR-Cas9-mediated deletion of the genomic region containing rs12928014. Data are shown as mean ± SEM from three independent experiments. P-value was calculated by two-sided t-test. P < 0.05 was deemed significant. (j) Luciferase reporter assay of the rs12928014. *Left*, schematic of the mini-promoter pGL6 construct and rs12928014-mini-promoter pGL6 luciferase reporter vectors. *Right*, luciferase activity in HEK293T cells (n = 3 independent experiments). Data are shown as mean ± SEM. P-values were cacualted using one-way ANOVA. Significance levels are indicated as *p < 0.05, **p < 0.01, ***p < 0.001, and ****p < 0.0001.

### Integration of SLE and eGFR GWAS with eQTL data reveals potential causal genes for LN

We next prioritized genes likely contributing to LN susceptibility by integrating Chinese SLE GWAS [31] and East Asian eGFR GWAS [32] with LN whole-blood eQTL(cf) data using Bayesian colocalization [33] **(Fig 5a)**. This prioritized 23 high-fidelity causal genes for SLE and 14 for kidney function **(Figs 5f, 5g, and Supplementary Fig S15, Supplementary Tables S19 and S20)**, including known SLE-associated genes (*PRKCB*, *BLK*, *FAM167A*) [34], Asian risk genes (*B3GALT6*, *STIM1*) [35], kidney injury markers (*SPP1* [36], *PAX8* [37]), and novel candidates (*INPP5B*, *PPP2R3C*, *FHL3*) **(Supplementary Fig S16)**. Interestingly, *PRKCB, INPP5B*, *ACOX1*, and *SPP1,* correlated with fibrosis in 433 micro-dissected human kidney tubules, with *ACOX1, SPP1*, and *PRKCB* also correlating with eGFR **(Fig 5h, Supplementary Fig S17, and Supplementary Table S21)**. At the *PRKCB* locus, centered around the SLE GWAS-prioritized SNP rs12928014 (P_GWAS_ = 2.5e-04), where GWAS associations and gene expression changes likely share the same causal variants in LN patient blood **(Fig 5g)**. LN patients with a higher dosage of the G allele showed increased *PRKCB* expression **(Fig 5b)**. ScRNA-seq showed *PRKCB* enrichment in B Mem **(Supplementary Fig S18)**. CRISPR-Cas9 deletion of the rs12928014-containing fragment in Raji B cells reduced *PRKCB* expression, and the G allele enhanced expression in dual-luciferase reporter assays **(Figs 5i and** **5j**).

Using the Drug Gene Interaction Database [38], 15 of 37 prioritized LN causal genes are druggable with approved indications, including *PRKCB* (diabetic neuropathy), *BLK*, *STIM1* and *ADK* (antineoplastic), and *SPP1* (acne and immunosuppressant) **(Supplementary Table S22 and S23)**. Cell-type profiling revealed 6 colocalized kidney function genes were highly expressed in neutrophils **(Supplementary Fig S21)**, whereas SLE colocalized genes formed five clusters (C1 to C5), with most HLA genes enriched in B cells (C5) **(Supplementary Fig S20 and Supplementary Table S24)**. Major histocompatibility complex (MHC) region require further validation due to its complex genetic architecture.

### Linking fine-mapped SNPs to CREs reveals cell-type regulatory programs in lupus nephritis

We integrated fine-mapped GWAS variants [39–41] with single-cell peak-to-gene linkages to access how causal SNPs disrupt CREs in LN phenotypic manifestations, including SLE and eGFR **(Fig 6a)**. Summing the posterior probabilities of fine-mapped SNPs overlapping gene-linked CREs, we identified 234 SLE- and 411 eGFR-linked genes (fmGWAS-linked genes) [42] **(Fig 6b and Supplementary Tables S25 and S26)**, with 31 shared **(Supplementary Fig S21, and Supplementary Table S27)**. Notably, *BLK* and *FAM167A* were also prioritized by colocalization **(Supplementary Table S19 and Fig 5g)**. Among 411 eGFR fmGWAS-linked genes, 10 were key LN-related TFs (e.g., *SPI1*) and 93 overlapped with differentially expressed HRGs **(Fig 6b)**. Similarly, among the 234 SLE fmGWAS-linked genes, 51 were differentially expressed HRGs and 8 were key TFs driving immune dysregulation. Shared genes were enriched for metabolic process **(Supplementary Table S28 and Supplementary Fig S22a)**, whereas eGFR-specific fmGWAS-linked genes were enriched for myeloid apoptosis and fatty acid metabolic processes **(Supplementary Table S29 and Supplementary Fig S22b)**, and SLE-specific fmGWAS-linked genes for B cell proliferation, and T cell mediated cytotoxicity **(Supplementary Table S30 and Supplementary Fig S22c)**. Both gene sets were primarily expressed in myeloid cells, with additional B-cell enrichment for SLE fmGWAS-linked genes **(Figs 6c and** **6d**). Interestingly, *ETS1*-linked CREs harbored causal SNPs of both traits **(Figs 6e-6g, Supplementary Table S25 and S26)** and were more accessible in lymphoid cells, whereas eGFR fine-mapped variants overlapped myeloid-specific CREs linked to *TET2*, and casual SLE SNPs overlapped *BACH2*-linked CREs highly accessible in BIN.

**Figure 6.**
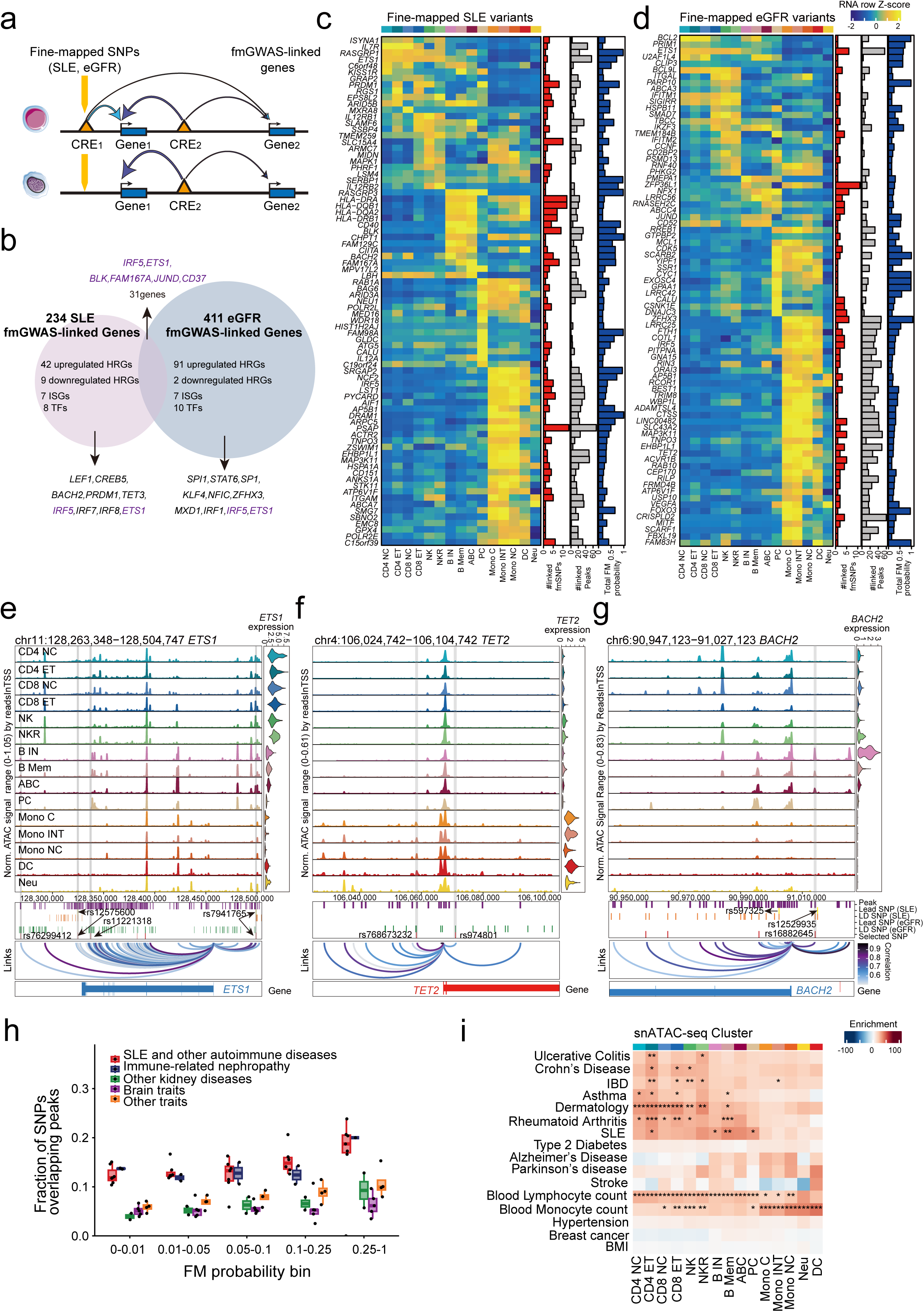
Integration of fine-mapped GWAS variants with single-cell peak-to-gene linkages reveals key genes and cell types contributing to lupus nephritis. (a) Experimental scheme. Fine-mapped GWAS variants were integrated with single-cell peak-to-gene linkages to assess their disruption of CREs and contribution to LN phenotypic manifestations (SLE[41] and eGFR[39, 50]). Genes linked to fine-mapped variants (fmGWAS-linked genes) were identified by summing the posterior probabilities of SNPs overlapping linked CREs. (b) This analysis identified 234 SLE-fmGWAS-linked genes and 411 eGFR-fmGWAS-linked genes, 31 of which are shared. Among the SLE-fmGWAS-linked genes, 51 are previously defined differentially expressed HRGs in LN PBMCs (42 upregulated, 9 downregulated), 8 are TFs and 7 are ISGs implicated in immune dysregulation in LN. Among the eGFR-fmGWAS-linked genes, 10 TFs and 7 ISGs are implicated in LN, and 93 overlap with these differentially expressed HRGs (91 upregulated, 2 downregulated). (c) The top 80 genes linked to peaks containing fine-mapped SNPs for SLE. The heatmap shows relative gene expression across high-resolution scRNA clusters, with cell types color-coded in the top bar. The red bar plot to the right indicates the number of linked fine-mapped SNPs per gene, the blue bar plot shows the sum of fine-mapped posterior probability for these SNPs, and the gray bar plot represents the total number of peak-to-gene linkages identified for each gene. (d) The top 80 genes linked to peaks containing fine-mapped SNPs for eGFR. The heatmap shows relative gene expression across high-resolution scRNA clusters, with cell types color-coded in the top bar. The red bar plot to the right indicates the number of linked fine-mapped SNPs per gene, the blue bar plot shows the sum of fine-mapped posterior probability for these SNPs, and the gray bar plot represents the total number of peak-to-gene linkages identified for each gene. (e) Genomic tracks for chromatin accessibility at the *ETS1* locus. Right: violin plots showing integrated *ETS1* expression across cell types. Below (*top* to *bottom*): pseudobulk ATAC-seq peaks from the entire PBMC dataset, lead SNPs and their LD proxies from fine-mapped SLE and eGFR GWAS loci, selected SNPs, and peak-to-gene linkages (loop) identified in our dataset. Gray vertical bars across both panels highlight selected fine-mapped SNPs overlapping CREs linked to *ETS1* expression. (f) Genomic tracks for chromatin accessibility at the *TET2* locus. Right: violin plots showing integrated *TET2* expression across cell types. Below (*top* to *bottom*): pseudobulk ATAC-seq peaks from the entire PBMC dataset, lead SNPs and their LD proxies from fine-mapped SLE and eGFR GWAS loci, selected SNPs, and peak-to-gene linkages (loop) identified in our dataset. Gray vertical bars across both panels highlight selected fine-mapped SNPs overlapping CREs linked to *TET2* expression. (g) Genomic tracks for chromatin accessibility at the *BACH2* locus. Right: violin plots showing integrated *BACH2* expression across cell types. Below (*top* to *bottom*): pseudobulk ATAC-seq peaks from the entire PBMC dataset, lead SNPs and their LD proxies from fine-mapped SLE and eGFR GWAS loci, selected SNPs, and peak-to-gene linkages (loop) identified in our dataset. Gray vertical bars across both panels highlight selected fine-mapped SNPs overlapping CREs linked to *BACH2* expression. (h) Fraction of fine-mapped (FM) SNPs overlapping cell type-specific open chromatin regions, binned by increasing fine-mapping posterior probability. Each dot represents on GWAS trait, with boxplot colors indicating trait groups (see **Supplementary Table S8** for details). The center lines show the medians; the box limits indicate the 25th and 75th percentiles; the whiskers extend to the 5th and 95th percentiles; the outliers are represented by the dots. (i) LDSC identifies enrichment of GWAS SNPs for autoimmune and non-autoimmune traits in open chromatin regions across LN PBMC immune cell types. Asterisks indicate the FDR-adjusted significance levels: *FDR□<□0.05, **FDR□<□0.005, ***FDR□<□0.0005.

### Genetic evidence implicates B cells in autoimmune kidney traits and CD4 effector T cells in LN treatment response

To prioritize driver cell types for autoimmune disorders and kidney traits, we ranked and binned fine-mapped SNPs by their posterior probability for GWAS traits **(Supplementary Note and Supplementary Table S31)**. Fine-mapped SNPs for autoimmune disorders and immune-mediated nephropathies, such as IgA nephropathy and membranous nephropathy, were preferentially enriched in immune cell open chromation, with enrichment increasing for higher-confidence SNPs **(Fig 6h, Supplementary Fig S23 and Supplementary Table S31)**. SLE fine-mapped SNPs were enriched in B Mem and ABCs **(Supplementary Fig S23 and Supplementary Table S32)**. Intriguingly, prioritized casual SNPs for membranous nephropathy also showed ABC enrichment, supporting a shared B cell-driven mechanism. Fine-mapped variants associated with the response to cyclophosphamide, an established frontline treatment of LN, were enriched in CD4^+^ effector T cells, suggesting a potential role for these cells in influencing LN treatment outcomes **(Supplementary Fig S23 and Supplementary Table S32)**.

We next performed stratified linkage disequilibrium score regression (LDSC) [43] to integrate cell-type-specific open chromatin with GWAS data for 16 traits **(Supplementary Table S33)**. GWAS signals for SLE and other immune-mediated disorders, such as ulcerative colitis, Crohn’s disease and inflammatory bowel diseases, were most enriched in CD4^+^ effector T cells **(Fig 6i and Supplementary Table S34)**. GWAS signals for SLE, asthma, dermatology, and rheumatoid arithritis were consistently enriched in B cells, pariticually in B Mem **(Fig 6i and Supplementary Table S34)**.

### TF-driven regulatory programs underlying B cell differentiation in LN

Given the strong enrichment of peak-to-gene linkages and autoimmune heritability in B cells **(Fig. 5e and** **6i****, Supplementary Fig S9a and S20a)**, we examined B cell subset dynamics. LN PBMCs showed pronounced PC accumulation and increased expression of key PC transcription regulators and markers **(Fig. 7a, 1e, Supplementary Figs S24 and S25a, and Supplementary Table S35)**, with upregulated genes enriched for endoplasmic reticulum (ER) targeting and IFN-γ response **(Supplementary Fig 25b and Supplementary Table S36)**, and downregulated immunoglobulin production genes **(Supplementary Table S37),** indicating proliferating PCs with reduced antibody synthesis amid elevated ER stress and immune activation.

**Figure 7.**
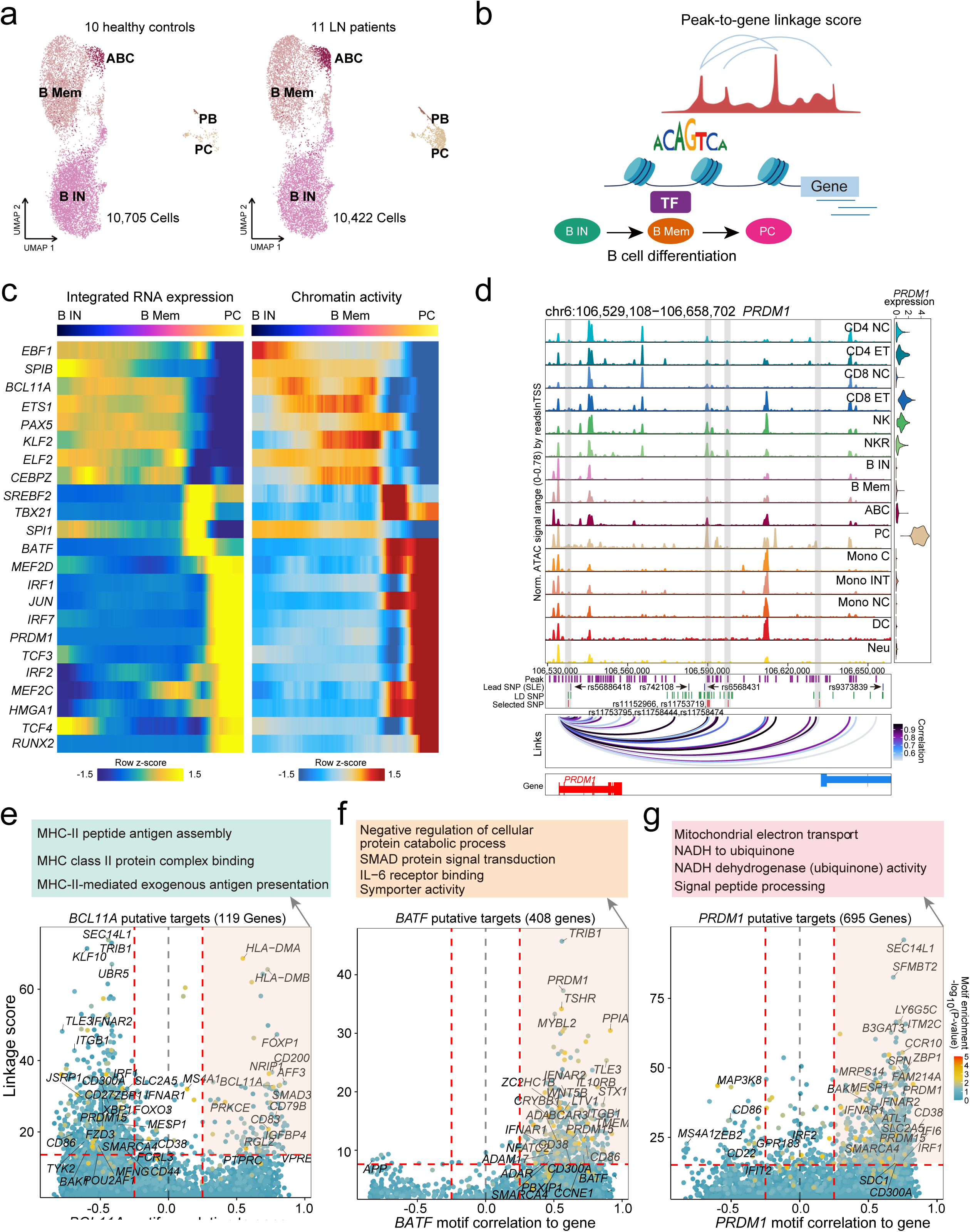
Integrative analysis of transcriptional regulation underlying B cell subset alterations in lupus nephritis. (a) UMAP visualization of B cells from healthy controls (n = 10) and LN patients (n□=□11), with individual cells colored by annotated subclusters. B IN: naïve B cells, B Mem: memory B cells, ABC: age-associated B cells, PB: plasmablasts, PC: plasma cells. Each dot represents an individual cell. (b) Experimental scheme. TF activity-gene expression correlations were integrated with linkage scores quantifying the confidence of TF binding motifs within associated CREs. Target genes were defined as those showing both strong expression correlation with global motif activity and high linkage score, identifying genes likely regulated by CRE-bound TFs. (c) Heatmaps showing TF regulators correlated with B cell differentiation along pseudotime. The left panel shows TF motif activity, while the right panel displays corresponding TF expression. Both datasets are Z-score-scaled across cell types per TF for visualization. Only TFs with consistent significant correlations between motif activity and expression are shown. Genomic tracks for chromatin accessibility at the *PRDM1* locus. Right: violin plots showing integrated *PRDM1* expression across cell types. Below (*top* to *bottom*): pseudobulk ATAC-seq peaks from the entire PBMC dataset, lead SNPs and their LD proxies from fine-mapped SLE GWAS loci, selected SNPs, and peak-to-gene linkages (loop) identified in our dataset. Gray vertical bars across both panels highlight selected fine-mapped SNPs overlapping CREs linked to *PRDM1* expression. (d) Prioritization of *BCL11A* gene targets. (e) Prioritization of *BATF* gene targets. (f) Prioritization of *PRDM1* gene targets. The x-axis represents Pearson’s correlation between TF motif activity and integrated gene expression across all expressed genes across B cells. The y-axis indicates the TF linkage score, calculated as the sum of motif score scaled by peak-to-gene link correlation for all linked peaks. Color of points denotes the hypergeometric enrichment of the TF motif within all linked peaks for the corresponding gene. Top gene targets are indicated in the shaded area (motif correlation to gene expression > 0.25, linkage score > 80th percentile). Functional annotation (gene ontology) of the top targets is shown above.

Next, we identified key TFs defining B cell subsets by analyzing differential motif accessibility and correlating TF expression with motif activaty, revealing known regulators of B cell identity **(Supplementary Fig S26)**. Using a semi-supervised pseudotemporal trajectory across B IN, B Mem, and PCs, we observed sequential, stage-dependent TF activation **(Fig 7b and7c, and Supplementary Fig S27)**, with several TFs also linked to fine-mapped GWAS variants, including *PRDM1* **(Fig 7d, Supplementary Table S25 and S26)**. TF regulatory targets were inferred by integrating TF activity-gene expression correlations with CRE motif confidence **(Supplementary Note**, **Fig 7b)**. We highlighted three representative TFs to illustrate stage-specific regulatory networks during B cell differentiation. *BCL11A* (B IN) regulated 119 putative targets enriched for MHC-II antigen processing and presentation **(Fig 7e, Supplementary Tables S37 and S38)**; *BATF* (B Mem) controlled JAK/STAT signaling and type 1 IFN signaling **(Fig 7f, Supplementary Tables S38 and S39)**; and *PRDM1* (PCs) regulated type I IFN response, signal peptide processing and mitochondrial electron transport pathways **(Figs 7e, 7g, Supplementary Tables S38 and S39)**, with over half of its targets negatively correlating with *BCL11A*. Stage-dependent chromatin accessibility and expression confirmed their regulatory roles in B cell differentiation **(Supplementary Figs S28a-c).**

### Single-cell regulatory map implicates SLE GWAS mechanisms, regulatory regions, genes, and cell types

Finally, we explored whether our single-cell regulatory atlas could resolve cell types and target genes underlying LN-associated GWAS signals. Most SLE and eGFR GWAS variants lie in non-coding regions, making cell type-specific regulatory context essential for functional interpretation. Integrating our single-cell regulatory atlas with SLE [31] and eGFR [32] GWAS data identified 841 and 353□ variants overlapping CREs, regulating 260 SLE- and 191 eGFR-associated genes, respectively **(Supplementary Tables S40 and S41)**.

As an example, we examined the *FAM167A* locus, repeatedly associated with SLE in multiple GWAS [44, 45]. On chromosome 8, SLE-associated variants were also likely causal for altered *FAM167A* expression in LN blood **(Fig 8a)**. Specifically, the SLE-associated variant rs4840568 (*P*_GWAS_ = 3.61e-10) [31] influenced *FAM167A* expression **(Supplementary Fig S29a)** and resided within a B cell-specific CRE between the *BLK* and *FAM167A* promoters **(Fig 8b)**. Peak-to-gene linkage analysis of the entire PBMC dataset revealed interactions between *FAM167A* open chromations and CREs harboring rs4840568 and other significant GWAS SNPs. Gapped *k*-mer support vector machine-based methods (gkm-SVM and deltaSVM) [46, 47] predicted a strong regulatory effect, with higher TF binding affinity for the A allele in BIN **(Fig 8c)**, consistent with higher *FAM167A* expression in A allele carriers **(Supplementary Fig S29a)**.

**Figure 8.**
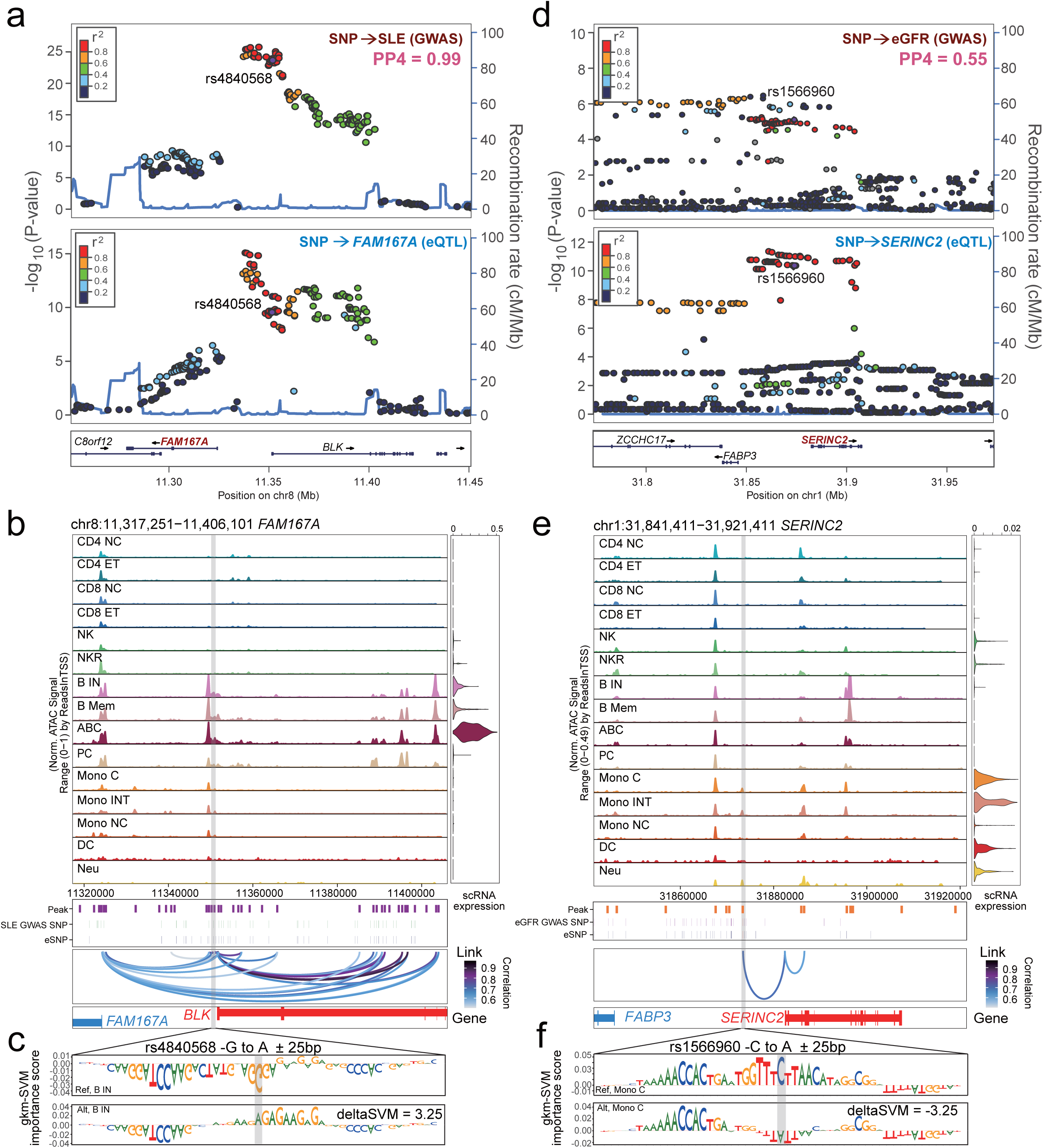
Integrative single-cell regulatory map provides insights into GWAS mechanisms linking regulatory regions, genes, and cell types. (a) LocusZoom plots of SLE GWAS variants (*top*) and LN blood *FAM167A* eQTL(cf)s (*bottom*). The x-axis shows the ±100 kb genomic region around rs4840568. The y-axis represents the significance -log_10_(two-sided P-value). Each data point represents a variant and the color represents the r^2^ (the degree of LD). n = 12,653 individuals for SLE GWAS; n = 99 individuals for LN blood eQTL(cf)s. (b) Genomic tracks for chromatin accessibility at the *FAM167A* locus. Right: violin plots showing integrated *FAM167A* expression across cell types. Below (*top* to *bottom*): pseudobulk ATAC-seq peaks from the entire PBMC dataset, GWAS significant SNPs (P-value < 5e-5) and eSNPs, and peak-to-gene linkages (loop) identified in our dataset. Gray vertical bars across both panels highlight selected GWAS SNP overlapping cell type specifc CREs linked to *FAM167A* expression. (c) Gkm-SVM method-estimated[46] transcription factor binding activity score for each base within ±25-base pair (bp) region surrounding rs4840568 for the reference (G) and alternative (A) alleles from the gkm-SVM model corresponding to theB cells. The SNP of interest is highlighted in grey. (d) LocusZoom plots of eGFR GWAS variants (*top*) and LN blood *SERINC2* eQTL(cf)s (*bottom*). The x-axis shows the ±100 kb genomic region around rs1566960. The y-axis represents the significance -log_10_(two-sided P-value). Each data point represents a variant and the color represents the r^2^ (the degree of LD). n = 297,355 individuals for eGFR GWAS; n = 99 individuals for LN blood eQTL(cf)s. (e) Genomic tracks for chromatin accessibility at the *SERINC2* locus. Right: violin plots showing integrated *SERINC2* expression across cell types. Below (*top* to *bottom*): pseudobulk ATAC-seq peaks from the entire PBMC dataset, GWAS significant SNPs (P-value < 5e-5) and eSNPs, and peak-to-gene linkages (loop) identified in our dataset. Gray vertical bars across both panels highlight selected GWAS SNP overlapping cell type specifc CREs linked to *SERINC2* expression. (f) Gkm-SVM method-estimated[46] transcription factor binding activity score for each base within ±25-base pair (bp) region surrounding rs1566960 for the reference (C) and alternative (A) alleles from the gkm-SVM model corresponding to the classical monocytes. The SNP of interest is highlighted in grey.

Another example is the *SERINC2* region on chromosome 1, centered around the eGFR-associated SNP rs1566960 (P_GWAS_ = 8.25e-06) [32], where GWAS associations and gene expression changes likely share the same causal variants in LN patient blood **(Fig 8d)**. eQTL analysis confirmed a strong association between rs1566960 and *SERINC2* expression **(Supplementary Fig S29b)**. This SNP resides in myeloid-specific accessible chromatin **(Fig 8e)**. Peak-to-gene linkage analysis of the entire PBMC dataset further revealed strong correlation between open chromatin regions harboring rs1566960 (and other GWAS SNPs) and *SERINC2*. DeltaSVM analysis revealed that TF binding activity within a ±25 bp window around rs1566960 decreased when the allele changed from C to A in classical monocytes cells **(Fig 8f)**. Correspondingly, LN patients with a higher dosage of the A allele showed decreased *SERINC2* expression **(Supplementary Fig S29b)**.

## Discussion

In this study, we established a comprehensive single-cell multi-omics atlas of PBMCs from newly diagnosed, minimally treated LN patients and applied orthogonal analytical frameworks to prioritize key TFs, causal genes, and disease-relevant cell types. Integrated scRNA-seq and snATAC-seq analyses revealed extensive immune circuit rewiring, marked by heightened innate immune activation and impaired adaptive immunity in LN. We defined critical CRE-gene regulatory modules, with peak-to-gene linkages strongly enriched in B cells, and identified TFs orchestrating these programs. To extend these insights to a broader disease population, we generated a blood eQTL atlas from 99 Chinese LN patients, prioritizing 14 likely causal genes for kidney function and 23 for SLE. Fine-mapped SNP-to-CRE mapping highlighted critical cell type convergence of autoimmune disorders and immune-mediated nephropathies, with B cells showing strong heritability enrichment for both SLE and membranous nephropathy. Further dissection of TF-directed programs in B cell subsets revealed differentiation stage specific regulatory networks governed by *PRDM1*, *BCL11A*, and *BATF*. Finally, we clarified how eGFR- and SLE- associated GWAS variants exert their regulatory effects within distinct single-cell chromatin contexts.

By focusing on newly diagnosed, minimally treated LN patients, we captured immune alterations intrinsic to LN rather than secondary to therapy, addressing a key limitation of prior single-cell studies largely conducted in heterogenous SLE cohorts. Integrating scRNA-seq and snATAC-seq from the same patients, we generated the first single-cell regulatory atlas of PBMCs, revealing coordinated epigenetic and transcriptional reprogramming driving impaired adaptive immunity and enhanced innate inflammatory activation in LN.

We further established a blood eQTL resource for a Chinese LN cohort, enabling population-specific genotype-expression mapping and Bayesian colocalization that prioritized 14 high-fidelity causal genes for kidney function and 23 for SLE. Notably, *PRKCB*, previously implicated in autoantibody production and immune complex-mediated kidney injury [48], was shown to be regulated by the variant rs12928014 in B cells through eQTL mapping, allele-specific reporter assays, and CRISPR-mediated perturbations.

This study is limited by its focus on circulating PBMCs, which may not fully capture kidney-resident immune cells or the complex microenvironments within the kidney. Future studies integrating longitudinal blood sampling with kidney biopsy-derived single-cell data will further clarify regulatory dynamics during disease progression and treatment.

In conclusion, we reconstruct the transcriptional regulatory architecture of immune dysregulation, providing a framework to dissect disease mechanisms, prioritize causal genes and TFs, and guide precision therapeutic strategies.

## Data Availability

All data produced in the present study are available upon reasonable request to the authors.

## Acknowledgements

This work is supported by the National Natural Science Foundation of China [32300534, 32270659 to X.S., and 82100765 to T. C.], Basic Research Program of Jiangsu [BK20243061 to Z. L.], the Open Project of Jiangsu Provincial Science and Technology Resources (Clinical Resources) Coordination Service Platform [TC2023006 to X.S.], the Key R&D Program of Zhejiang Province [2024SSYS0021 to X.S.], the Natural Science Foundation of Zhejiang Province [LZ25C060002 to X.S. and LQ24C060001 to J.Z.], the Fundamental Research Funds for the Central Universities [K20250114 to X.S.], and the hundred talents program of Zhejiang University to X.S.; We thank the technical support from the Core Facilities, Liangzhu Laboratory, Zhejiang University.

## KEY MESSAGES

### What is already known on this topic

- Recent studies have advanced understanding of lupus nephritis (LN) by defining immune cell heterogeneity and pathogenic cell states, including extrafollicular B cells, cytotoxic CD8⁺ T cells, and VCAM1⁺ proximal tubule cells.
- Genome-wide association studies (GWAS) have identified numerous genetic risk loci for SLE and LN; however, how these variants influence immune cell states and regulatory programs in patients remains poorly understood.
- Most existing studies have focused on transcriptional signatures of LN, leaving the regulatory architecture uderlyling immune dysregulation and its genetic determinants insufficiently characterised.

### What this study adds

- This study provides an integrated single-cell multi-omics atlas of peripheral blood mononuclear cells (PBMCs) from newly diagnosed, minimally treated LN patients, revealing extensive immune remodelling characterized by heightened innate immune activation and impaired adaptive immune responses orchestrated by cell-type-specific transcription factor (TF) centered regulatory circuits.
- We generated a blood eQTL atlas from 99 Chinese LN patients, prioritizing 14 high-fidelity causal genes for kidney function and 23 for SLE, and confirmed correlations between several of these genes and kidney function and fibrosis in 433 micro-dissected human kidney tubule samples.
- Integration with fine-mapped SLE GWAS variants highlighted strong heritability enrichment in B cells, where TF-driven programs define stage-specific differentiation networks.

### How this study might affect research, practice or policy

- This work provides a systems-level framework linking genetic risk to cell-type-specific regulatory mechanisms in LN, complementing existing kidney-focused single-cell studies.
- Identification of TF-centred regulatory circuits and genetically prioritized causal genes offers mechanistic targets for future functional studies and therapeutic development.
- Our integrative multi-omics and genetic approach supports the development of precision medicine strategies for LN, enabling patient stratification and treatment design beyond histopathology-based classification.

## Author contributions

1. X. S., Z. L., and H. Z. conceived, planned, and oversaw the study and wrote the manuscript. H. Z. and F. Y. analyzed data with the help of X. S., T. C., X. L., K. S., and Z. L. T. C. and F. Y. performed the sample collection. H. Z. performed the CRISPR-Cas9 mediated genome editing with the help of J. Z., S. C., and Z. M., J. Z., N. L., X. Y. X.S., H. Z., T. C., F. Y., and Z. L. assisted with data generation and manuscript revision.

## Data availability

The human PBMC scRNA-seq, snATAC-seq, and whole blood RNA-seq datasets, and genotyping microarray data, and whole blood eQTL data are available upon reasonable request to us. Predicted super-enhancer-associated genes [19] across 86□human cell types and tissues are available as supplemental data 1 of Hnisz *et al* [19]. Enhancer-gene interactions from the ABC model [17] of 131 human tissues and cell types are downloaded from the website (https://www.engreitzlab.org/resources/). Fine-mapped SNPs for 22 GWAS traits were obtained from two sources: (1) a compendium of fine-mapped SNPs for 94 UK Biobank traits (www.finucanelab.org/data)[39], from which we selected traits including eGFR, height, body mass index (BMI), and systolic blood pressure (SBP); and (2) pre-computed fine-mapped SNPs generated by PICS2 [40, 41] (Probabilistic Identification of Casual SNPs), which infers the probability of causality for variants within a locus based on GWAS signal strength and local linkage disequilibrium. These PICS fine-mapped datasets were obtained from the GWAS catalog (https://pics2.ucsf.edu/Downloads/PICS2-GWAScat-2021-06-11.txt.gz) [40, 41]. Formatted summary statistics for LDSC were downloaded from the LDSC website (https://console.cloud.google.com/storage/browser/broad-alkesgroup-public-requester-pays/sumstats_formatted).

## Competing interest statement

The authors declare no competing interests.

## Notes

### Competing Interest Statement

The authors have declared no competing interest.

