## Supplementary Information for "Regulatory architecture underlying immune dysregulation reconstructed by single-cell multi-omics in lupus nephritis"

**This PDF file includes:**

Supplementary Note

Figs. S1 to S31

### Supplementary Note

### Methods

#### Human PBMC scRNA-seq data generation

Fresh blood was collected from 10 healthy donors and 11 LN patients. Peripheral blood mononuclear cells (PBMCs) were isolated by dilluting the samples 1:2 in phosphate-buffered saline (PBS; devoid of $Ca^{2+}$ and $Mg^{2+}$) supplemented with 2% fetal bovine serum (FBS) and 2 mM EDTA. A 35 mL aliquot of the diluted sample was transferred to 50 mL Falcon tubes and carefully overlaid ont 15 mL of Ficoll-Paque PLUS (GE Healthcare, 17-1440-02). Density gradient centrifugation was performed at 400 × g for 35 minutes without brake. The PBMC layer was carefully harvested and washed twice with PBS before proceeding to single-cell RNA-sequencing library preparation.

Single-cell RNA-seq libraries were prepared using the Single Cell 3' Library and Gel Bead Kit v3.1 (10x Genomics, 1000121) together with the Chromium Single Cell G Chip Kit (10x Genomics, 1000120). Cell suspensions (300-600 viable cells/μL, assessed by Countstar) were loaded onto the Chromium Single Cell Controller following the manufacturer's protocol. Cells were suspended in PBS containing 0.04% BSA, and approximately 6,000 cells were loaded per channel, targeting a recovery of ~3,000 cells. Captured cells were lysed, and RNA was barcoded via reverse transcription within individual Gel Beads-in-Emulsion (GEMs). Reverse transcription was performed on a Bio-Rad S1000™ Touch Thermal Cycler at 53°C for 45 min, followed by 85°C for 5 minutes, and then held at 4°C. The resulting cDNA was amplified and assessed using an Agilent 4200 system. Sequencing libraries were prepared according to the manufacturer's instructions and sequenced on an Illumina NovaSeq 6000 platform using a paired-end 150 bp (PE150) strategy, with a minimum depth of 100,000 reads per cell.

Raw scRNA-seq data (fastq files) were processed using Cell Ranger (v6.1.2, 10x Genomics) for read alignment to the hg19 reference genome (GRCh37), barcode demultiplexing, and feature-barcode matrix generation.

Downstream quality control, normalization, and dimensionality reduction were performed using R package Seurat (v4.4.0) [1]. Cells with fewer than 400 detected genes or more than 2,500 genes were excluded to remove low-quality cells and potential doublets. Cells with mitochondrial gene content exceeding 20% were also removed to eliminate potential artifacts from damaged cells.

After quality control, the scRNA-seq count matrix of 237,864 high-quality PBMCs was normalized using the *LogNormalize* function implemented in R package Seurat (v4.4.0) [1], with a scale factor of 1E04. Batch effects were adjusted using Harmony (v1.2.0) [2] **(Supplementary Fig S30)**. Dimensionality reduction was performed using Principal Component Analysis (PCA), and the top 15 Harmony-adjusted principal compents (PCs) were selected to generate a *k*-nearest neighbor graph. Clustering was then performed using the Louvain algorithm [3] at a resolution of 0.8. Uniform Manifold Approximation and Projection (UMAP[4], v0.2.9.0) was applied for visualization. Cell clusters were annotated based on previously established marker genes [5, 6], and clusters with fewer than 100 cells were excluded from further analysis. Genes expressed in fewer than five cells were discarded to reduce noise.

**Interferon and inflammatory score estimation**

LN patients were stratified by disease severity using the Activity Index (AI) [7], a histopathologic scoring system that captures active renal inflammation with greater specificity in renal pathology than the SLEDAI-2K [8]. Patients with AI scores ≤ 7 were assigned to the moderate group, while those with scores > 7 were considered severe. To evaluate gene set activity, we applied AUCell (v1.21.1) [9] to the scRNA-seq expression matrix, focusing on previously reported SLE-related interferon-stimulated genes (ISGs) [10] and inflammatory gene signatures[11] **(Supplementary Table S6)**. Genes expressed in fewer than 1% of cells were removed, and for each cell, expression values of the retained genes were ranked. *AUCell_buildRankings* and *AUCell_calcAUC* functions were then used to compute area under the curve (AUC) scores for each gene set. Statistical testing was carried out within each cell types using the Wilcoxon Rank Sum test, and two-sided P-values were adjusted for multiple comparisons with the Benjamini-Hochberg (BH) correction.

**Differentially expressed gene identification**

For differential expression analysis between health controls and LN patients, raw gene counts were summed at the sample level to generate gene-by-sample matrices. Genes with ≤ 30 read counts in fewer than three samples were excluded, and library sizes were normalized using the TMM (trimmed mean of M-values) method implemented in the *calcNormFactors* function to adjust for sequencing depth differences. Differential expression was evaluated using the *exactTest* function implemented in the R package edgeR (v3.36.0) [12]. Genes were defined as **differentially expressed if they met both** a false discovery rate (FDR) < 0.05 and **an absolute $\log_2$ fold change ($|\log_2 FC|$) > 0.5.**

**Cell-cell communication analysis**

Intercellular communication was examined using Cellchat [13] (v1.6.1) to infer signaling networks from single-cell transcriptome data. Significant cell-cell interactions were identified based on overexpressed ligand-receptor pairs within each cell population ($p < 0.05$). Differences in incoming and outgoing signaling strength across all cell clusters were then quantified between healthy controls and LN paitents. Directional signaling dynamics were further assessed by comparing communication probabilities of specific ligand-receptor pairs between defined sender and receiver populations using the *netVisual_bubble* function with the *compare* parameter activated.

**Human PBMC snATAC-seq data generation**

Nuclei were isolated from PBMCs and prepared for single-nucleus ATAC-seq following the 10x Genomics protocol, *Nuclei Isolation for Single Cell ATAC*

*Sequencing* (CG000169). Extracted nuclei were washed and resuspended in chilled Diluted Nuclei Buffer (10x Genomics, 2000153), with the volume adjusted according to the initial cell count and target nuclei concentration. Nuclei were counted using the Countstar Rigel S2 and immediately processed for library preparation. Using the 10x Genomics Single Cell ATAC Solution, nuclei were encapsulated into nanoliter-scale GEMs with the Chromium Chip E Single Cell Kit (10x Genomics, 1000156) and the Chromium Single Cell ATAC Library & Gel Bead Kit (10x Genomics, 1000110). Approximately 750,000 unique 10x barcodes were used to individually index transposed DNA from each nucleus. Libraries were prepared following the manufacturer's instructions and sequenced on an Illumina NovaSeq platform using a paired-end 50 bp (PE50) strategy, achieving a minimum depth of 25,000 read pairs per nucleus.

Raw snATAC-seq data were processed using Cell Ranger ATAC [14] (v2.1.0) for read alignment, deduplication, and transposase cut site identification, with sequencing reads aligned to the hg19 (GRCh37) reference genome. Quality control was performed with ArchR (v1.0.2) [15], calculating unique fragments and transcription start site (TSS) enrichment per cell to assess signal-to-noise ratio. True cells were distinguished from low-quality droplets via expectation-maximization clustering using mclust (v6.1) [16] on log10-transformed fragment counts (nFragments) and TSS enrichment values (Mclust(df, G=2:4, modelNames=VVV)), retaining cells from the distribution with the highest mean TSS enrichment **(Supplementary Fig S31)**. Cells with TSS enrichment scores below 5 or fewer than 1,000 fragments were additionally removed. Of 11 LN PBMC samples, 2 were excluded for insufficient true cells, leaving 9 for downstream analyses. Potential doublets were filtered using *addDoubletScores* and *filterDoublets* functions with a filter ratio of 1.

Dimensionality reduction and batch effect removal were performed using iterative latent semantic indexing (LSI) with 15,000 variable features and 15 dimensions, implemented in ArchR via the *addIterativeLSI* function, **(Supplementary Fig S31)**. Cells were clustered using the *addClusters* function at a resolution of 0.4, and two-dimensional embeddings were generated using the *addUMAP* function with parameters nNeighbors = 35, minDist = 0.3, and metric = "cosine". Clusters were manually annotated to PBMC cell types based on a published list of marker genes [5, 6], consistent with the assignments used in scRNA-seq data. For visualization, gene activity scores were smoothed using the MAGIC algorithm [17] to improve interpretability.

For each annotated cell type, reproducible peak sets were identified using the *addReproduciblePeakSet* function in ArchR [15], which calls MACS2 [18] (v2.1.2) with the parameters --nomodel --nolambda --shift -75 --extsize 150. Peaks were required in at least 25 cells, capped at 500 peaks per cell and 150,000 peaks per cluster. Peaks were extended ±250 bp from summits, and mitochondrial (chrM) and Y chromosome peaks were excluded. Significant peaks were defined by q-value ≤ 0.1, with reproducibility across pseudo-bulk replicates (minReplicates = 2) enforced. Differentially accessible regions (DARs) were identified from the PeakMatrix using the *getMarkerFeatures* function in ArchR [15], grouping cells by cluster. Technical covariates such as TSS enrichment and fragment counts were regressed out, and differential accessibility was assessed using a Wilcoxon rank-sum test. DARs were considered significant at FDR ≤ 0.01 and |log2 fold change| ≥ 1.

**Motif enrichment with ChromVAR**

TF motif enrichment analysis was performed using chromVAR (v.1.16.0) [19], integrated within the ArchR framework. We applied the *addMotifAnnotations* function, mapping CIS-BP [20] motifs onto the consensus peak set, thereby annotating potential TF binding sites, or onto DARs to identify motifs enriched in specific cell types. To account for technical and sequence biases, background peak sets were generated with the *addBgdPeaks* function, matched for genomic features such as GC content, copy number, and accessibility. Subsequently, we used the *addDeviationsMatrix* function to compute motif deviation z-scores for each cisBP motif, quantifying cell-to-cell variability in motif accessibility and thereby inferring TF activity across single cells.

#### Integration of scRNA-seq and snATAC-seq data

Integration of scRNA-seq and snATAC-seq data was performed using the *addGeneIntegrationMatrix* function in ArchR with 3,000 highly variable genes. Cross-modality anchors were identified using the *FindTransferAnchors* function [1] implemented in Seurat, which applies canonical correlation analysis (CCA) to match each snATAC-seq cell to its most similar scRNA-seq cell. Gene activity scores for snATAC-seq were calculated by summing fragments within gene bodies and 2 kb upstream promoter regions and then normalized. Cluster-level averages of scRNA-seq expression and snATAC-seq gene activity were computed, and Z-scores were calculated for each gene. Pearson's correlation across the top 3,000 shared variable genes confirmed strong concordance between transcriptomic and epigenome cluster assignments **(Fig. 2d)**.

#### Peak-to-gene linkages identification and validation

Potential cis-regulatory elements (CREs) were linked to target genes using the *addPeak2GeneLinks* function implemented in ArchR. This procedure involved the creation of up to 500 partially overlapping pseudobulks, and each composed of 100,000 nearest-neighbor single cells, referred to as "low-overlapping cell aggregates." For each pseudobulk, chromatin accessibility (peak counts) and integrated scRNA-seq expression counts were summed. Peaks located within ±250 kb of a gene transcription start site (TSS) were paired with that gene, and Pearson's correlation coefficients were calculated between $\log_2$-normalized peak accessibility and gene expression. Associations with correlation coefficients greater than 0.5 were considered high-confidence. To improve sensitivity for closely related subtypes, the analysis was repeated within each major cell type using subcluster-specific peaks [21]. Peak-to-gene links from both the entire and subclustered datasets were merged into a consensus set, duplicates were removed, and links were ranked by correlation strength, resulting in 91,497 high-confidence associations **(Supplementary Fig S9a)**.

Linkages were validated by comparison to enhancer-gene interactions from the ABC model [22] (131 human tissues and cell types; https://www.engreitzlab.org/resources/). A linkage was considered validated if the peak overlapped an ABC-defined enhancer and the associated gene matched. Enrichment of ABC-predicted links was tested against all possible peak-gene pairs within 250 kb. To account for distance bias, a distance-matched background was generated by binning peak-to-gene distances into 20 equal groups and sampling matched sets. Enrichment was assessed using a hypergeometric test over 100 iterations.

**Highly regulated gene (HRG) identification**

HRGs were defined by ranking all expressed genes according to the number of peak-to-gene links. Genes associated with at least 20 peaks, corresponding to the inflection point of the distribution, were designated as HRGs, yielding a total of 1,178 genes **(Fig. 3b)**. These HRGs were compared with previously reported super-enhancer-associated genes across multiple tissues and cell lines, and enrichment significance was evaluated using a hypergeometric test **(Fig. 3c)**. For functional annotation, the top 250 HRGs from each k-means cluster were used for GO enrichment analysis with R package topGO (v2.46.0) [23] using the *weight01* method, which accounts for the GO hierarchy. Following topGO guidelines, no additional multiple hypothesis testing correction was applied.

**Pseudobulk profiling of chromatin accessibility and gene expression**

Pseudobulk profiles were generated using the function *getLowOverlapAggregates* implemented in ArchR, grouping 250 *k*-nearest neighbor cells with a maximum overlap of 80%. For each pseudobulk, aggregated matrices were computed for chromatin accessibility (PeakMatrix), gene activity (GeneScoreMatrix), and integrated expression (GeneIntegrationMatrix). Peak counts were summed across cells, normalized to 10,000 reads per pseudobulk, and $log_2$-transformed. GeneScore and GeneIntegration values were averaged across cells and $log_2$-transformed for downstream analyses.

For heatmap visualization of peak accessibility, log-transformed counts were quantile-normalized using the *preprocessCore* package (v1.61.0) and converted to z-scores across pseudobulks. Peaks linked to the same gene were hierarchically clustered using Euclidean distance and complete linkage, with co-accessible peak clusters defined via a tree-cut threshold (typically 1-5 clusters per gene). Pseudobulks were labeled according to the predominant cell-type annotation among constitute cells.

To examine the relationship between chromatin accessibility and gene expression, total gene accessibility was calculated as the $log_2$ sum of depth-normalized counts across linked peaks, and compared with $log_2$-transformed integrated expression per pseudobulk. Correlations between GeneAcitivity scores and integrated expression were also assessed. Results were visualized using scatter plots with linear regression fits and heatmaps displaying gene-linked peak accessibility alongside pseudobulk-level expression values.

**TF regulatory network inference**

To identify transcriptional regulators driving expression variations of HRGs and ISGs, we used FigR package (v0.1.0) [24] to combine motif enrichment with correlations between TF expression and chromatin accessibility. Each gene was grouped with its *k*-nearest neighbors (*k* = 30) based on accessibility profiles of linked CREs. Motif enrichment analysis was performed on CREs within each gene group relative to matched background peaks. For each TF, RNA expression values were smoothed across similar cells to reduce technical noise and dropout effects, and then correlated with the aggregate accessibility of its target genes using Spearman's correlation. To quantify TF regulatory potential, evidence from motif enrichment and expression-accessibility correlation was integrated into a composite regulation score. A signed probability score, termed the regulation probability, was derived from Z-test P-values of both analyses and transformed into a $-\log_{10}$ regulation score for interpretability. Regulatory networks of selected TFs and target genes were visualized using R package networkD3 (v0.4).

**B cell subclustering analysis and trajectory inference**

After annotating major immune lineages, B cell populations, including naïve, memory, age-associated B cells, and plasma cells, were subsetted for downstream analysis. For the scRNA-seq data, the Seurat and Harmony workflows were largely reapplied. The top 2,000 highly variable genes were selected, scaled, and analyzed by PCA. Reclustering was performed on the first 13 Harmony-adjusted PCs at a resolution of 2, yielding transcriptionally distinct B cell subsets annotated based on the expression of known marker genes [5, 6]. For the snATA-seq data, B cells were processed using iterative LSI on 25 dimensions with 25,000 variable features. Batch effects were corrected with Harmony [2], followed by graph-based clustering (resolution = 0.4) and UMAP visualization (nNeighbors = 35, minDist = 0.4, metric = "cosine"). Clusters with fewer than 100 cells were excluded. Gene activity scores were then integrated with matched scRNA-seq data using the *addGeneIntegrationMatrix* function, enabling refined annotation of B cell subsets.

A B cell differentiation trajectory was inferred using the *addTrajectories* function on UMAP embeddings, including only naïve, memory, and plasma cells, with the BIN cluster designated as the root. Candidate transcriptional regulators were identified by correlating chromVAR motif deviation z-scores with TF expression across aggregated B cell groups and by analyzing integrated gene expression and chromVAR motif deviation scores along the trajectory using the c*orrelateTrajectories* function to evaluate relationships between TF motif activity and gene expression during B cell differentiation.

Potential TF target genes were defined as previously described [21]. Pearson's correlation coefficients were computed between chromVAR motif activity of candidate TFs and expression of all genes. For each TF-gene pair, a linkage score was calculated by summing the products of squared peak-to-gene correlations and motif scores for all linked peaks containing the TF motif, prioritizing genes supported by multiple strongly correlated peaks enriched for high-confidence TF motifs.

**Genotype and RNA-seq data production**

Genotyping was performed for 101 LN patients using the Infinium Asian Screening Array-24 v1.0 BeadChip (Illumina). Quality control steps were conducted in PLINK [25] (v1.90). Two samples were excluded due to sex inconsistencies between RNA-seq and genotype data. Monomorphic SNPs and SNPs with a minor allele frequency (MAF) $< 0.05$, Hardy-Weinberg equilibrium $P < 1e-6$, and call rate $< 90\%$ were excluded. SNPs located on sex chromosomes were excluded. After quality control, 99 individuals and 293,143 SNPs remained for downstream analysis **(Supplementary Table S42)**. Principal component analysis (PCA) was then performed using EIGENSTRAT [26] on the genotype data of 99 participants together with 2,504 individuals from the 1,000 Genomes Project Phase 3 [27] (NCBI build 37, release date October 2014). An additional 333 SNPs absent or with incompatible alleles in the 1,000 Genomes dataset were removed. Genotype data were subsequently pre-phased with SHAPEIT2 [28] and imputed with IMPUTE2 [29] using the multiethnic panel reference from 1,000 Genome Phase 3. Post-imputation filters excluded SNPs with MAF $< 5\%$, Hardy-Weinberg equilibrium $p < 1E-06$, missing rate $< 95\%$ for best-estimated genotypes at posterior probability $> 0.9$, and imputation confidence score, INFO (a measure of $r^2$) $< 0.4$ (estimated by SNPTEST [30]) were excluded. As a result, 4,833,457 high-quality autosomal SNPs were retained for the eQTL analysis.

Total RNA was extracted from whole blood samples of the same 101 LN patients and assessed using the Agilent 5400 TapeStation. Libraries were prepared with the Fast RNA-seq Lib Prep Kit V2 (ABclonal, RK20306), including mRNA capture, fragmentation, strand-specific cDNA synthesis, adapter ligation, and amplification. Library quality and concentration were evaluated using Qubit3.0 (Invitrogen) and qRT-PCR (BIO-RAD, T100), then sequenced on NovaSeq X Plus. The RNA-seq data consisted of strand-specific paired-end 150 bp reads prepared using the dUTP method, which preserves strand orientation by incorporating dUTP during second-strand cDNA synthesis. Sequence quality was assessed with *FastQC* function in trim_galore (v0.6.10), adapter sequences and low-quality bases were trimmed with Trim-Galore (v0.6.10). RNA-seq reads were aligned to the hg19 (GRCh37) reference genome [31] with STAR (v2.7.10) [32]. BAM files were coordinate-sorted and duplicates marked using samtools (v1.16.1) [33]. Gene-level read counts were quantified with RSEM (v1.3.1) [34]. Outliers were identified through hierarchical clustering based on average and cosine distances of the gene expression matrix. A two-sided $\mathcal{X}^2$ P-value was cacluated from the Mahalanobis distance in a 20-dimensional PC space. No outliers were detected. Sex compatibility was assessed based on X- and Y-chromosomal gene expression, and two samples with inconsist sex were removed from downstream analysis. After quality control, 99 LN patients with high quality RNA-seq, genotype data, and complete phenotype records were retained. Read counts were normalized across samples using the TMM method implemented in edgeR (v3.36.0) [35], and gene expression levels were estimated as transcripts per million (TPM). Only genes with TPM ≥ 0.1 in at least 5% of samples were considered expressed and used for downstream analysis.

### eQTL mapping using cell fraction model

As our previous study demonstrated that inclusion of cell fractions in the general eQTL mapping model substantially increased the power to identify eGenes (i.e., genes associated with at least one SNP), we performed *cis*-eQTL (hereafter referred to as eQTL) mapping on 99 bloood samples from LN patients using the cell fraction eQTL(cf) model [36]. To estimate blood cell fractions, we used CIBERSORTx [37], a reference-guided deconvolution tool for bulk tissue transcriptomes, since our previous study [36] showed that CIBERSORTx outperformed other deconvolution methods. To minimize bias arised from differences in PBMC cell type proportions, we randomly down-sampled 700 cells per cell type (matching the least abundant cell type) and used the marker gene expression matrix from our human PBMC scRNA-seq dataset as the reference. Deconvolution was performed in absolute mode with at least 500 permutations. Common cell types and those with limited co-linearity were retained for downstream eQTL identification **(Supplementary Figs S12a and S12b)**.

We next estimated the probablilistic estimation of expression residuals (PEER) factors from gene expression data, adjusting for age, sex, top three genotype PCs, and estimated fractions for five selected cell types (including $CD4^+$ T, $CD8^+$ T, B cell, monocytes, and neutrophils). Ten PEER factors, which maximized the number of identifed eGenes **(Supplementary Fig S12c)**, were included in downstream eQTL identificaiton.

EQTL mapping was then performed using the cell fraction eQTL(cf) model[36], with inverse-normal transformed gene expression as the dependent variable and SNP dosage as the independent variable, adjusting for age, sex, top three genotype PCs, ten PEER factors, and the estimated cell fractions for the five selected cell types. Nominal P-values for SNP-gene pairs were cacluated using MatrixeQTL (v2.3) [38] with an additive linear regression mode. To identify significant eGenes, we applied FastQTL (v2.184) [39] with adaptive permutation ("--permute 10000"). Beta distributaion-adjusted empirical P-values were converted into q-values using Storey's *q* method implemented in R package qvalue (v2.26.0) [40], with $q \leq 0.05$ used as the significant threshold. To identify significant eVariants, an empirical cutoff ($P_t$), defined as the empirical

P-value of the gene closest to the 0.05 FDR threshold, was used to estimate the nominal P-value cutoff for each gene based on the beta distribution of the minimum P-value distribtuion $f(P_{min})$, obtained from its permutations.

**Colocalization analysis**

Colocalization analysis was performed with *coloc* (v5.2.2) [41] to estimate the posterior probabilities that SLE or eGFR GWAS signals share common causal variants with blood eQTLs identified by the eQTL(cf) model. Summary statistics from Asian SLE [42] and eGFR [43] GWAS were used, retaining variants with P-values $< 5e-5$. LD pruning ($r^2 \geq 0.8$) was performed with *swiss* (v1.0b4) (https://github.com/statgen/swiss) to avoid inflation from correlated GWAS variants representing the same signal. Variants in LD $r^2 \geq 0.8$ with the lead SNP at each locus were removed, and regions ±100 kb around pruned GWAS variants overlapping with eGenes were analyzed. In *coloc*, H3 tests the hypothesis that both traits (e.g., phenotype and gene expression) are associated but have indepdent causal variants, whereas H4 tests the hypothesis that both traits share the same causal variants (colocalization). Following previous studies [44], colocalization was defined as PP4 (posterior probability of H4) > 0.5. Results were visualized with LocusZoom (v0.4.8), and functional annotation was performed using DAVID (v2024q4) [45].

**RNA-seq data of human kidney samples**

Correlation analyses between gene expression and kidney structure damage (fibrosis) and kidney function (eGFR) were performed using a cohort of 433 micro-dissected human kidney tubular samples, as described in Sheng et al.[36] and herein referred to as the CKD cohort, whose demographic and clinical features are summarized in **Supplementary Table S21**.

**Integration of fine-mapped GWAS variants with snATAC-seq peaks to prioritize trait-associated genes**

To link fine-mapped variants to candidate genes for LN-related phenotypes (SLE[46] and eGFR [47, 48]), we adopted SNPs with posterior probability ≥ 0.01 that overlapped snATAC-seq peaks. For each gene, posterior probabilities of all SNPs within its associated peaks were summed. Genes with high cumulative probabilities or multiple peak associations were prioritized as putative trait-associated candidates **(Supplementary Tables S25 and S26)**. The top 80 genes, ranked by total fine-mapping probability, were visualized in a heatmap showing row-scaled expression across high-resolution scRNA-seq clusters, with the number of linked peaks and cumulative probabilities annotated alongside each gene **(Figs. 6c and 6d)**.

**Enrichment analysis of fine-mapped SNPs in open chromatin regions**

We assessed the enrichment of fine-mapped SNPs within cell type-specific open chromatin regions. Cell clusters with fewer than 40 cells were excluded due to insufficient power to identify reliable peaks, and only clusters with more than 5,000 peaks were retained. Fine-mapped SNPs were collected from 22 GWAS traits **(Supplementary Table S31)**, including SLE, IgA nehropathy, membranous nephropathy, and diabetic kidney disease, and 15 GWAS traits were first classified into five categories based on prior knowledge of diseases or trait biology **(Fig. 6h)**. For each trait, fine-mapped SNPs were ranked by posterior probability and grouped into bins representing increasing confidence levels. Enrichment of SNP bins within open chromatin peaks was assessed for each immune cell type. As a background, the full set of fine-mapped SNPs (posterior probability ≥ 0.01) aggregated across all traits, independent of peak overlap, was used. Enrichment was evaluated by comparing the number of SNPs overlapping peaks against this background set using a one-sided Fisher's exact test, with P-values adjusted for multiple testing using the Benjamini-Hochberg method.

**LDSC and partitioning SNP heritability**

We applied stratified LDSC (v.1.0.1) [49, 50] to estimate SNP heritability enrichment of autoimmune disorders, blood cell counts, hypertension, and other non-immune traits across cell type-specific DARs **(Supplementary Tables S33 and S34)**. Annotation files were generated from DARs for each cell type following official guidelines (https://github.com/bulik/ldsc/wiki). Partitioned heritability was calculated for 16 GWAS traits using the "ldsc.py" script, with the 1000 Genomes Project Phase 3 European ancestry reference panel and baseline v2.2 annotations. P-values were adjusted for multiple testing using the Benjamini-Hochberg method.

**Effects of genetic variants on chromatin accessibility**

To predict the impact of non-coding variants on chromatin accessibility, we extended each open chromation peak summit by ±250 bp. For each cell cluster, the top 20,000 most significant peaks (ranked by MACS2 q-value) were selected, and background sequences were generated using the *genNullSeqs* function in R package gkmSVM (v0.83) [51], matched for sequence length, GC content, and repeat fraction. Gkm-SVM (weighted gapped k-mer Support Vector Machine) is a sequence-based method for predicting and detecting the regulatory vocabulary encoded in the functional regions. Given the large training set, we applied LS-GKM[52], which implements advanced gapped k-mer based kernel functions, to train the SVM model for each cell cluster. Model performance was evaluated using 10-fold cross-validation, and all models achieved AUROC > 0.9, with the 'wgkm' kernel. Trained models were then applied with the *gkmpredict* function in LS-GKM to estimate the effects of 11 mers for each cell cluster. For each SNP identified in our eQTL analysis, 51 bp sequences centered on the variants (±25 bp) were constructed, and accessibility changes were quantified as deltaSVM scores using the "deltasvm.pl" script, which measures the predicted accessibility difference between the two alleles.

**Cell culture**

Raji, Jurkat, and HEK-293T cells were kindly provided by the laboratories of Dr. Xiaomin Yu, Dr. Jin Zhang, and Dr. Nan Liu (Zhejiang University). Raji and Jurkat cells were cultured in RPMI-1640 medium (Gibco, 8123416) supplemented with 10% FBS (Noverse, NFBS-2500A), and HEK-293T cells were cultured in DMEM (MeilunBio, MA0212) with 10% FBS. All cells were incubated at 37°C in a humidified atmosphere with 5% $CO_2$.

**Lentivirus production**

For lentiviral production, 5e5 HEK-293T cells per well were seeded into six-well plates and transfected the following day with 1.1μg lentiCas9-Blast (Addgene #52962), 495 ng pMD2.G (Addgene #12259), 910ng psPAX2 (Addgene #12260), and 9 μl Lipofectamine 2000 (Thermo Fisher, 11668-019). Six hours post-transfection, the medium was replaced with fresh culture medium. Viral supernatants were harvested 72 h later, clarified by centrifugation at 4°C for 10 min, aliquoted, and stored at -80°C.

**Lentiviral transduction and Cas9 selection**

Raji and Jurkat cells (5e5 cells/ml) were transduced with 100 μl lentiviral supernatant in the presence of 1 μl polybrene and 500 μl culture medium in 24-well plates. Untransduced cells were used as negative controls. After 24 h, medium was replaced, and 10 μg/ml blasticidin was applied for selection, refreshed every two days until all control cells died, enriching for Cas9-expressing cells.

**CRISPR-mediated enhancer deletion**

Dual-guide RNAs (gRNAs) flanking rs12928014 were designed using CHOPCHOP v3 (https://chopchop.cbu.uib.no/#) and synthesized by GenScript (Nanjing, China). Two gRNA pairs with high predicated efficiency and low off-target potential were selected **(Supplementary Table S43)**. Cas9-expressing

Raji cells (4e5) were electroporated with 48 pmol of each gRNA using the Lonza 4D-Nucleofector system (SG Cell Line 4D-Nucleofector™ X Kit V4XC-3032, program DS-104). Jurket cells (1e6) were electroporated with 120 pmol gRNA using the Lonza 4D-Nucleofector system (SE Cell Line 4D-Nucleofector Kit, V4XC-1032, program CL-120). After 72 h, knockout efficiency was evaluated by qPCR, and *PRKCB* expression was measured by RT-qPCR in samples with >30% deletion efficiency.

**Luciferase reporter assays**

The pGL6-CMV-Luc plasmid (Beyotime, D2091) was modified by removing the CMV promoter (KpnI/HindIII digestion) and inserting a 32 bp minipromoter amplified from pGL4.23 (Promega) using Uni Seamless Assembly (TransGen Biotech, R30830), generating minipromoter-pGL6-Luc. A 500 bp enhancer fragment containing rs12928014 (chr16:23,867,757-23,868,258, hg19) with either the A or G allele was amplified from genomic DNA and cloned into the KpnI site.

HEK-293T cells (5e4 per well) were seeded in white 24-well plates and transfected with 500 ng of either minipormoter-pGL6-Luc, rs12928014-A-minipromoter-pGL6-Luc, or rs12928014-G-minipromoter-pGL6-Luc along with 15 ng Renila luciferase control plasmid using Lipofectamine 2000 (Thermo Fisher, 11668-019). Luciferase activity was measured 72h post-transfection with the Dual-Glo Luciferase Assay System (Vazyme, DD1205). Primer sequences used for construct generation are listed in **Supplementary Table S44**.

**Supplementary Figure 1**

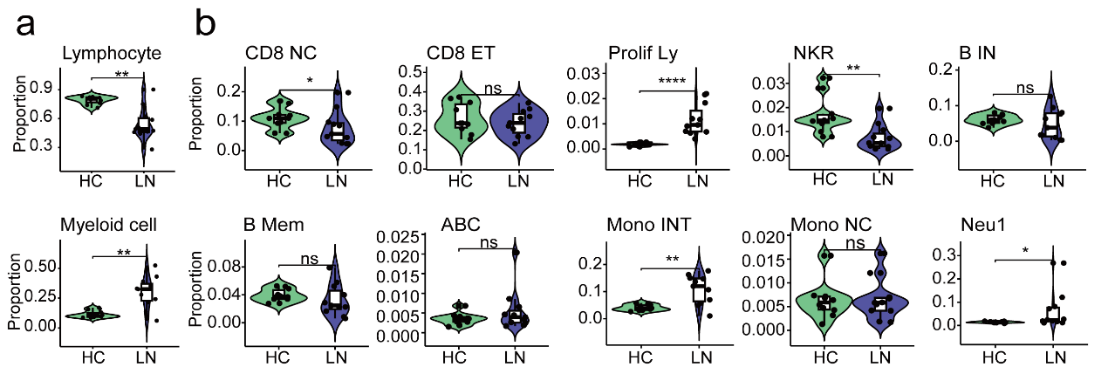


**Fig. S1. Comparison of cell fractions between healthy controls (HCs, n = 10) and LN patients (n = 11).** (a) Violin plots showing fractions of lymphocytes (*top*) and myeloid cells (*bottom*) in HCs and LN patients. (b) Violin plots showing cell fractions of their subpopulations. P-values were calculated using the Wilcoxon test comparing group means between LN and HC. Significance levels are indicated as *p < 0.05, **p < 0.01, ***p < 0.001, and ****p < 0.0001. CD8 NC: naïve $CD8^+$ T cells, CD8 ET: effector $CD8^+$ T cells, Prolif Ly: proliferating lymphocytes, NKR: natural killer recruiting cells, B IN: naïve B cells, B Mem: memory B cells, ABC: age associated B cells, Mono INT: intermediate ($CD14^{++}CD16^+$) monocytes, Mono NC: non-classical ($CD14^{dim}CD16^+$) monocytes, Neu1: neutrophil subpopulation 1.

### Supplementary Figure 2

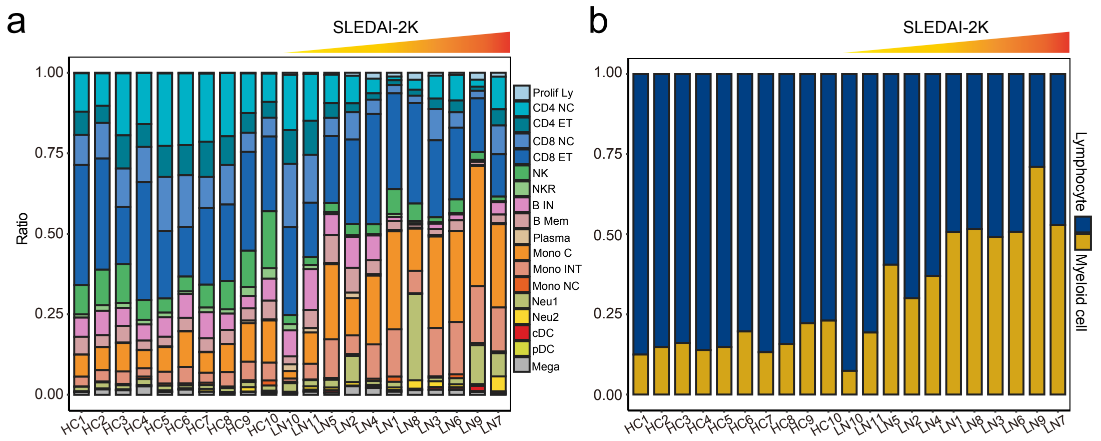


**Fig. S2. Bar plots showing relative cell abundances in scRNA-seq data from healthy controls (HCs, n = 10) and LN patients (n = 11).** Patients are ordered by increasing SLEDAI-2K scores. Colors represent (a) individual cell types and (b) two broad categories: lymphocytes and myeloid cells. CD4 NC: naïve $CD4^+$ T cells, CD4 ET: effector $CD4^+$ T cells, CD8 NC: naïve $CD8^+$ T cells, CD8 ET: effector $CD8^+$ T cells, Prolif Ly: proliferating lymphocytes, NK: natural killer cells, NKR: natural killer recruiting cells, B IN: naïve B cells, B Mem: memory B cells, ABC: age associated B cells, PC: plasma cells, Mono C: classical ($CD14^{++}CD16^-$) monocytes, Mono INT: intermediate ($CD14^{++}CD16^+$) monocytes, Mono NC: non-classical ($CD14^{dim}CD16^+$) monocytes, Neu1: neutrophil subpopulation 1, Neu2: neutrophil subpopulation 2, cDC: conventional dendritic cells, pDC: plasmacytoid dendritic cells, Mega: megakaryocytes.

**Supplementary figure 3**

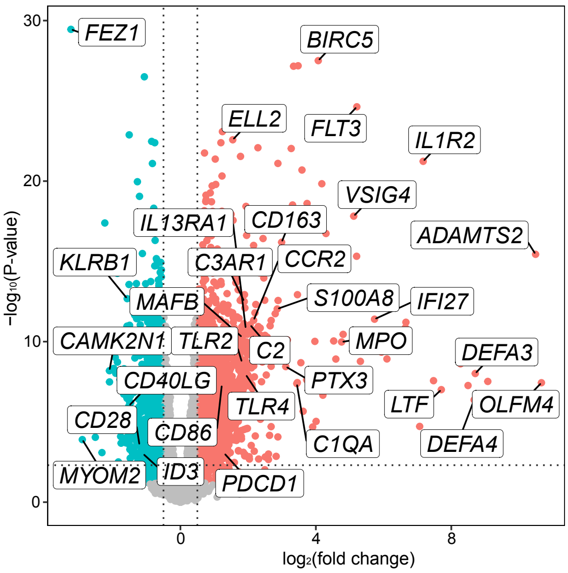


**Fig. S3. Volcano plot of differential gene expression from pseudobulk scRNA-seq data comparing LN patients (n = 11) and healthy controls (n = 10).** *X*-axis represents the base 2 log of the fold change for gene expression (read counts) between PBMCs of LN patients and healthy controls. *Y*-axis is the negative based 10 of the association P-value. A total of 1,939 upregulated genes in LN are shown in red color, and 1,285 downregulated genes are shown in light blue color. The horizontal dashed line marks - $\log_{10}$(P-value) = 1, and vertical dashed lines mark |$\log_2$ fold change| = 0.5. Each data point represents an individual gene.

### Supplementary figure 4

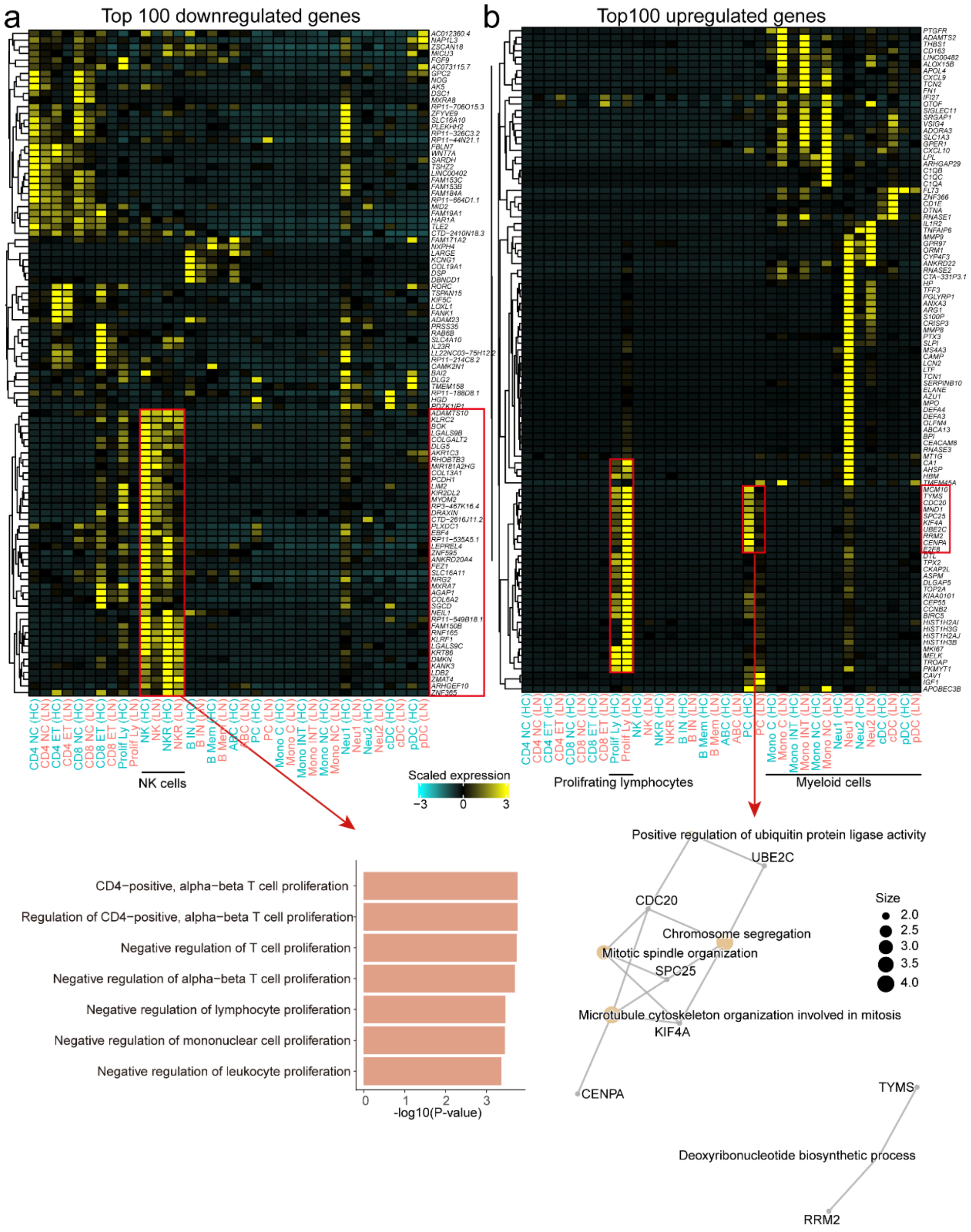


**Fig. S4. Expression patterns and functional annotation of the top 100 down- and upregulated DEGs across immune cell types in healthy controls (HCs) and LN patients.** (a) Top*:* Heatmap showing row-scaled expression of the top 100 downregulated genes in PBMCs of LN patients across 18 cell clusters (megakaryocytes excluded). The color scheme is based on Z-scores, calculated from normalized gene expression levels of the 18 cell types. Z-scores show the relatively expression levels compared to other clusters. Genes are hierarchically clustered based on expression profiles. Below: Functional annotation (gene ontology, GO) of 43 genes specifically expressed in NK cells among the top 100 downregulated DEGs. (b) same as in (a) but for the top 100 upregulated DEGs in PBMCs of LN patients. Below*:* Network visualization of the GO functional annotation for 10 genes upregulated at the pseudobulk level in LN but markedly downregulated in plasma cells, generated using the *cnetplot* function from R package clusterProfiler[53]. Nodes represent GO terms (orange) and DEGs (grey), with edges connecting genes to their enriched terms. Node size corresponds to $-\log_{10}$(P-value). CD4 NC: naïve $CD4^+$ T cells, CD4 ET: effector $CD4^+$ T cells, CD8 NC: naïve $CD8^+$ T cells, CD8 ET: effector $CD8^+$ T cells, Prolif Ly: proliferating lymphocytes, NK: natural killer cells, NKR: natural killer recruiting cells, B IN: naïve B cells, B Mem: memory B cells, ABC: age associated B cells, PC: plasma cells, Mono C: classical ($CD14^{++}CD16^{-}$) monocytes, Mono INT: intermediate ($CD14^{++}CD16^{+}$) monocytes, Mono NC: non-classical ($CD14^{dim}CD16^{+}$) monocytes, Neu1: neutrophil subpopulation 1, Neu2: neutrophil subpopulation 2, cDC: conventional dendritic cells, pDC: plasmacytoid dendritic cells.

### Supplementary figure 5

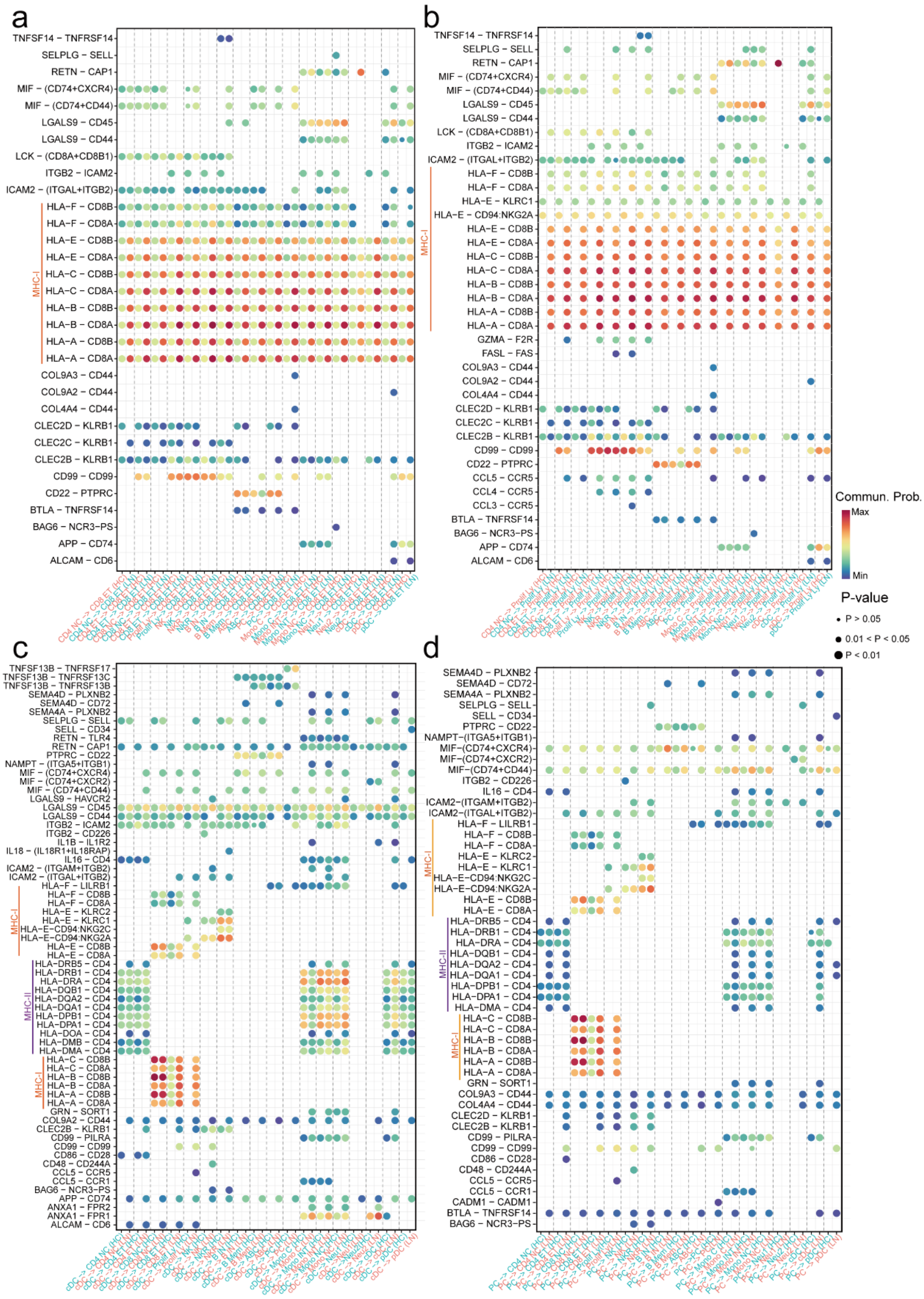


**Fig. S5. Significant ligand-receptor interactions mediating incoming signaling from other immune cell types to effector CD8⁺ T cells, proliferating lymphocytes, conventional dendritic cells, and plasma cells in healthy controls (HCs) and LN patients.** (a) Effector CD8⁺ T cells, (b)

proliferating lymphocytes, (c) conventional dendritic cells, (d) plasma cells. Dot color and size represent the calculated communication probability and p-values, respectively. P-values were computed using a one-sided permutation test. CD4 NC: naïve $CD4^+$ T cells, CD4 ET: effector $CD4^+$ T cells, CD8 NC: naïve $CD8^+$ T cells, CD8 ET: effector $CD8^+$ T cells, Prolif Ly: proliferating lymphocytes, NK: natural killer cells, NKR: natural killer recruiting cells, B IN: naïve B cells, B Mem: memory B cells, ABC: age associated B cells, PC: plasma cells, Mono C: classical ($CD14^{++}CD16^-$) monocytes, Mono INT: intermediate ($CD14^{++}CD16^+$) monocytes, Mono NC: non-classical ($CD14^{dim}CD16^+$) monocytes, Neu1: neutrophil subpopulation 1, Neu2: neutrophil subpopulation 2, cDC: conventional dendritic cells, pDC: plasmacytoid dendritic cells.

### Supplementary figure 6

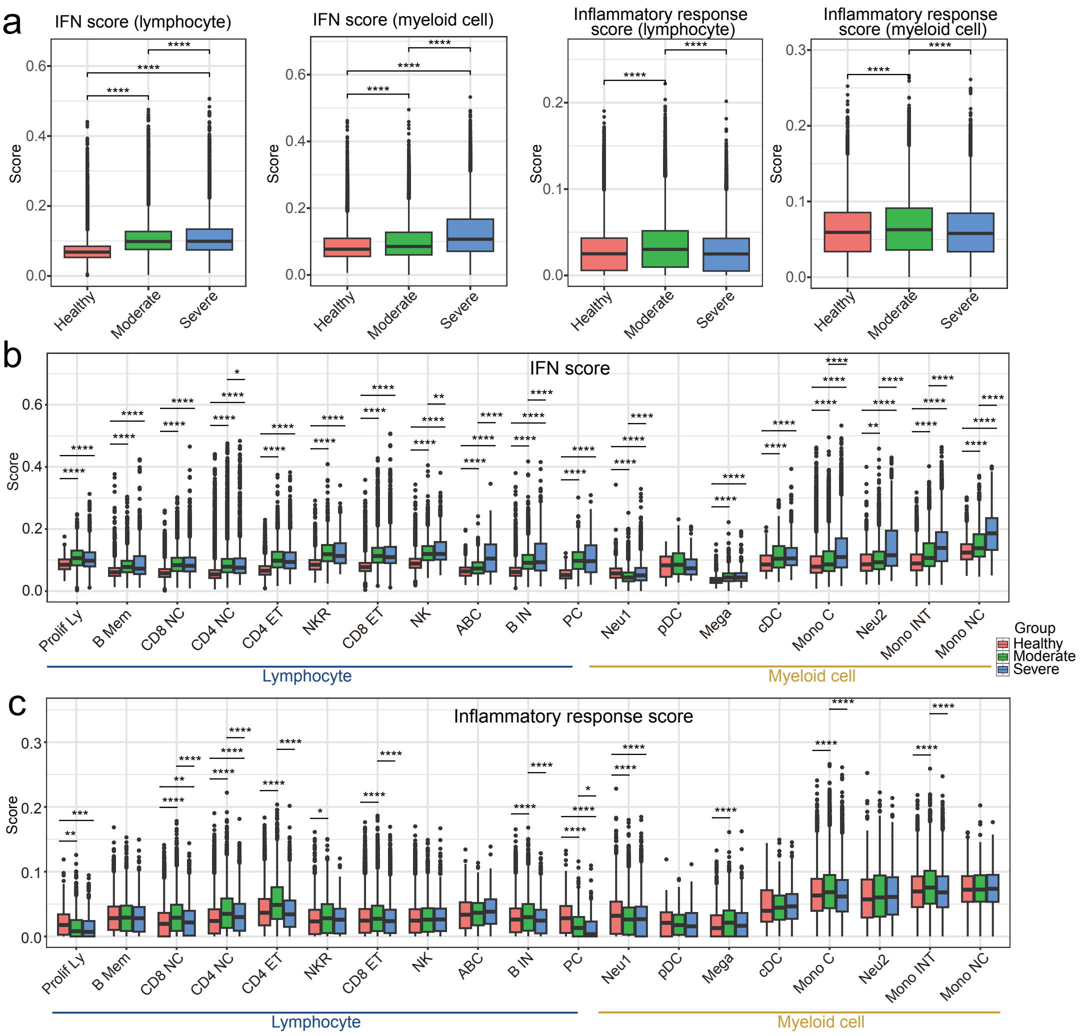


**Fig. S6. Interferon and inflammatory response scores across immune cell types in healthy controls (HCs) and LN patients at moderate and severe stages.** (a) Interferon (IFN) scores (*left*) and inflammatory response (IR) scores (*right*) in lymphocytes and myeloid cells from HC, moderate LN (AI ≤ 7), and severe LN (AI > 7). (b) IR scores and (c) IFN scores across immune cell types in HC, moderate LN, and severe LN. The center lines show the medians; the box limits indicate the 25th and 75th percentiles; the whiskers extend to the 5th and 95th percentiles; the outliers are represented by the dots. P-value was calculated using two-sided t-test with Bonferroni-adjusted *p*-values. Significance levels are indicated as *p < 0.05, **p < 0.01, ***p < 0.001, and ****p < 0.0001.

### Supplementary figure 7

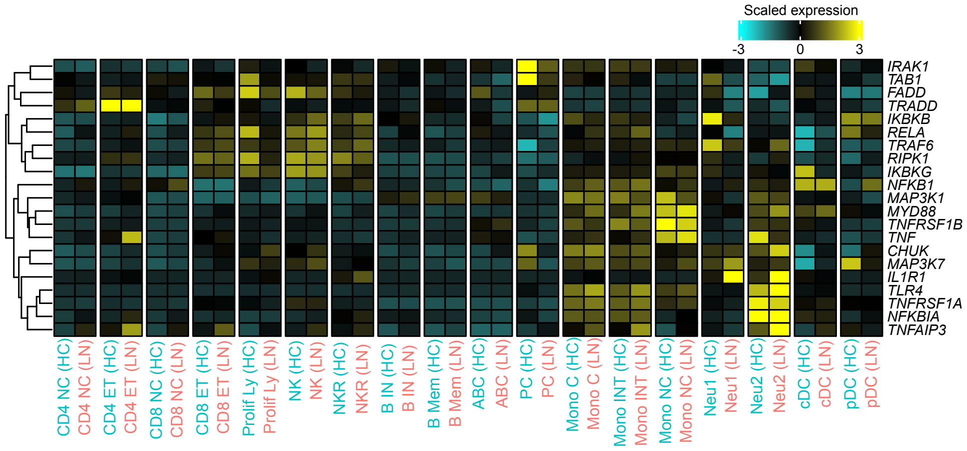


**Fig. S7. Expression patterns of 21 pro-inflammatory genes across immune cell types in healthy controls (HCs) and LN patients.** Heatmap showing row-scaled expression of the 21 pro-inflammatory genes across 18 cell clusters (megakaryocytes excluded). The color scheme is based on Z-scores, calculated from normalized gene expression levels of the 18 cell types. Z-scores show the relatively expression levels compared to other clusters. Genes are hierarchically clustered based on expression profiles. CD4 NC: naïve CD4$^+$ T cells, CD4 ET: effector CD4$^+$ T cells, CD8 NC: naïve CD8$^+$ T cells, CD8 ET: effector CD8$^+$ T cells, Prolif Ly: proliferating lymphocytes, NK: natural killer cells, NKR: natural killer recruiting cells, B IN: naïve B cells, B Mem: memory B cells, ABC: age associated B cells, PC: plasma cells, Mono C: classical (CD14$^{++}$CD16$^-$) monocytes, Mono INT: intermediate (CD14$^{++}$CD16$^+$) monocytes, Mono NC: non-classical (CD14$^{dim}$CD16$^+$) monocytes, Neu1: neutrophil subpopulation 1, Neu2: neutrophil subpopulation 2, cDC: conventional dendritic cells, pDC: plasmacytoid dendritic cells.

### Supplementary figure 8

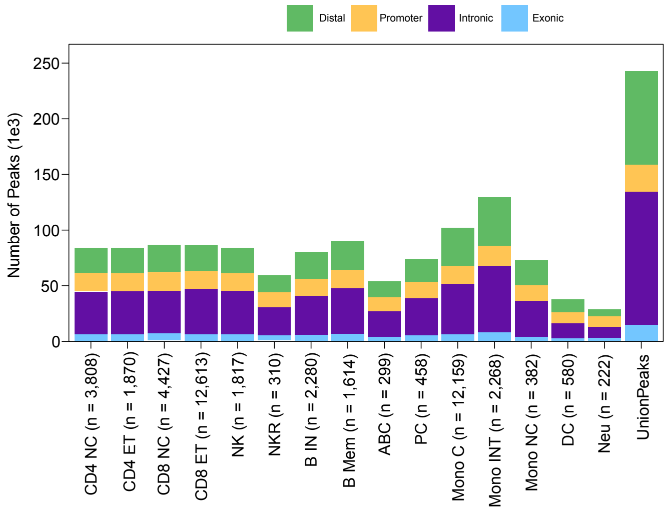


**Fig. S8. Bar plots showing the number of accessible peaks in distal elements, promoters, introns, and exons across immune cell types.** The color scheme is based on cell types. ABC: age associated B cells, B IN: naïve B cells, B Mem: memory B cells, CD4 ET: effector $CD4^+$ T cells, CD4 NC: naïve $CD4^+$ T cells, CD8 ET: effector $CD8^+$ T cells, CD8 NC: naïve $CD8^+$ T cells, DC: dendritic cells, Mono C: classical ($CD14^{++}CD16^-$) monocytes, Mono INT: intermediate ($CD14^{++}CD16^+$) monocytes, Mono NC: non-classical ($CD14^{dim}CD16^+$) monocytes, Neu: neutrophils, NK: natural killer cells, NKR: natural killer recruiting cells, PC: plasma cells, UnionPeaks: the union set of all accessible peaks across cell types.

### Supplementary figure 9

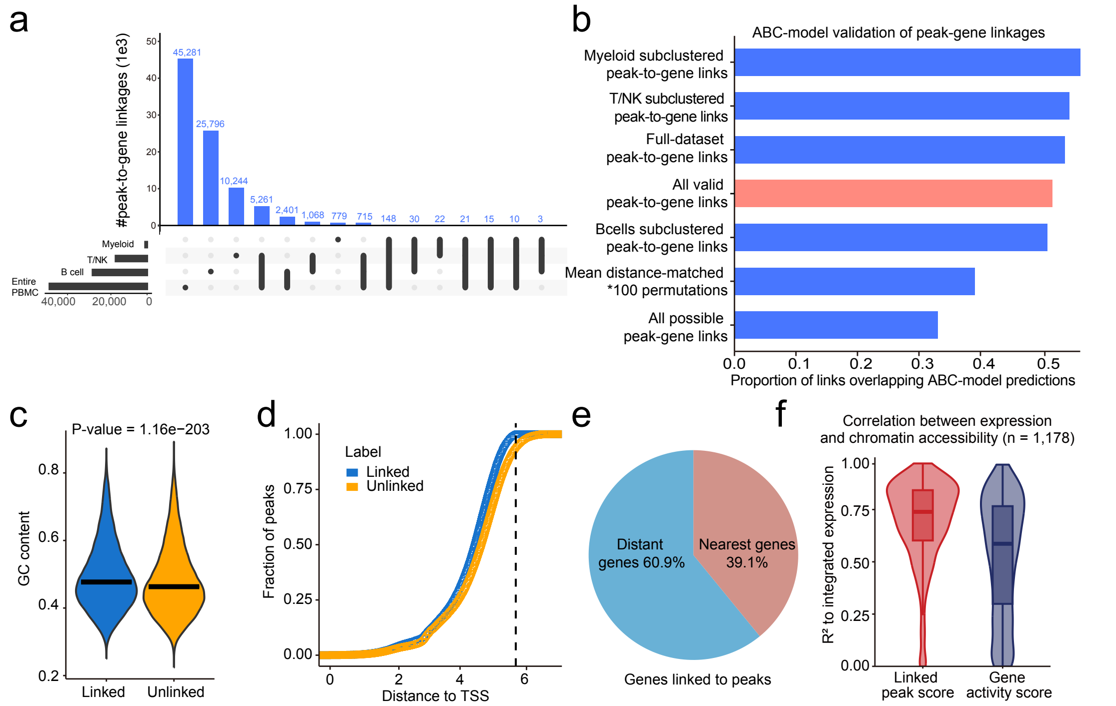


**Fig. S9. Characterization of peak-to-gene linkages across PBMC cell types.** (a) Number of shared and unique peaks among snATAC-seq cell types. Myeloid: the union set of accessible peaks from monocytes (including classical, non-classical and intermediate monocytes), dendritic cells, and neutrophils. T/NK: the union set of accessible peaks from naïve CD4$^+$ T cells, effector CD4$^+$ T cells, naïve CD8$^+$ T cells, effector CD8$^+$ T cells, natural killer cells, and natural killer recruiting cells. B cells: naïve B cells, memory B cells, age associated B cells, and plasma cells. Full Dataset: the entire PBMCs. (b) Bar plot showing the proportion of peak-to-gene linkages where both peak and gene were validated by a multi-tissue dataset [22, 54] of activity-by-contact (ABC) model enhancer-gene predictions. Categories compared included the space of all possible peak-to-gene links, the mean of 100 permutations drawn from all possible peak-to-gene links where for each permutation 146,088 peaks were selected to match the anchor distance distribution of true peak-to-gene links, and the set of true peak-to-gene links identified on each sub-clustered dataset. One-sided Fisher's exact test enrichment comparing each subgroup of true peak-to-gene links to a distance-matched background set, $p < 2.2e-16$. (c) Comparison of GC content between peaks with peak-to-gene associations and those without gene associations. The center lines show the medians; the box limits indicate the 25th and 75th percentiles; the whiskers extend to the 5th and 95th percentiles. Two-sided p-value was calculated using the Wilcoxon rank-sum test. (d) Cumulative distribution of peak fractions as a function of distance to the nearest TSS. Peaks with peak-to-gene associations (*blue*) and peaks without gene associations (*orange*) are shown. Distances are plotted on a $\log_{10}$ scale, and the vertical dashed line represents the maximum linkage distance (250 kb) used in the peak-to-gene association analysis. (e) Pie chart showing the relative proportions of peak-to-gene linkages linked to the nearest gene (*red*) and to more distant genes (*blue*). (f) Comparison of the linked peak score (sum of accessibility at linked peaks) compared to the gene activity score for predicting gene expression for the 1,178 HRGs. The center lines show the medians; the box limits indicate the 25th and 75th percentiles; the whiskers extend to the 5th and 95th percentiles.

### Supplementary figure 10

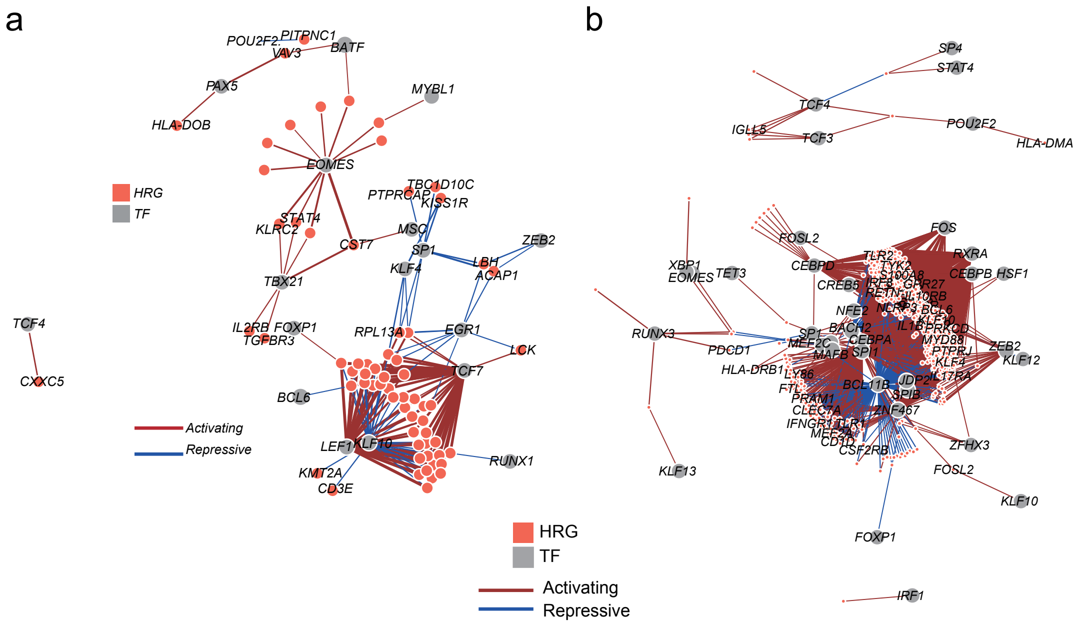


**Fig. S10. TF-gene regulatory networks underlying immune dysregulation in lupus nephritis.** (a) Network visualization of significant TF-regulated HRGs that are downregulated in LN PBMCs. (b) Network visualization of significant TF-regulated HRGs that are upregulated in LN PBMCs. Nodes represent FigR-defined target genes (*red*) and their associated TFs (*grey*). Edges are scaled and colored by the signed regulation score.

Supplementary figure 11

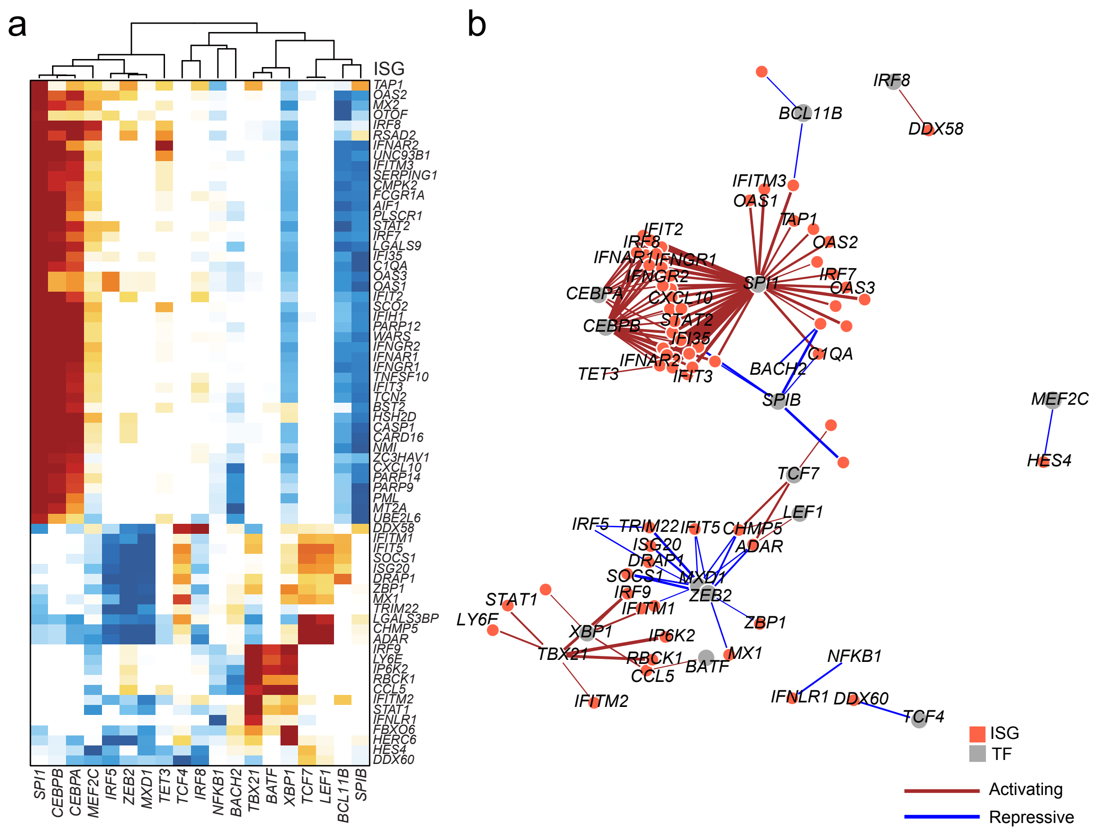


**Fig. S11. TF regulation of interferon-stimulated genes (ISGs).** (a) Heatmap showing regulation scores for TFs associated with ISGs. Positive and negative regulatory effects are shown in red and blue, respectively. Rows represent 68 target ISGs and columns represent 19 TFs. (b) Network visualization of significant TF-regulated ISGs. Nodes represent FigR-defined target genes (*red*) and their associated TFs (*grey*). Edges are scaled and colored by the signed regulation score.

**Supplementary Figure 12**

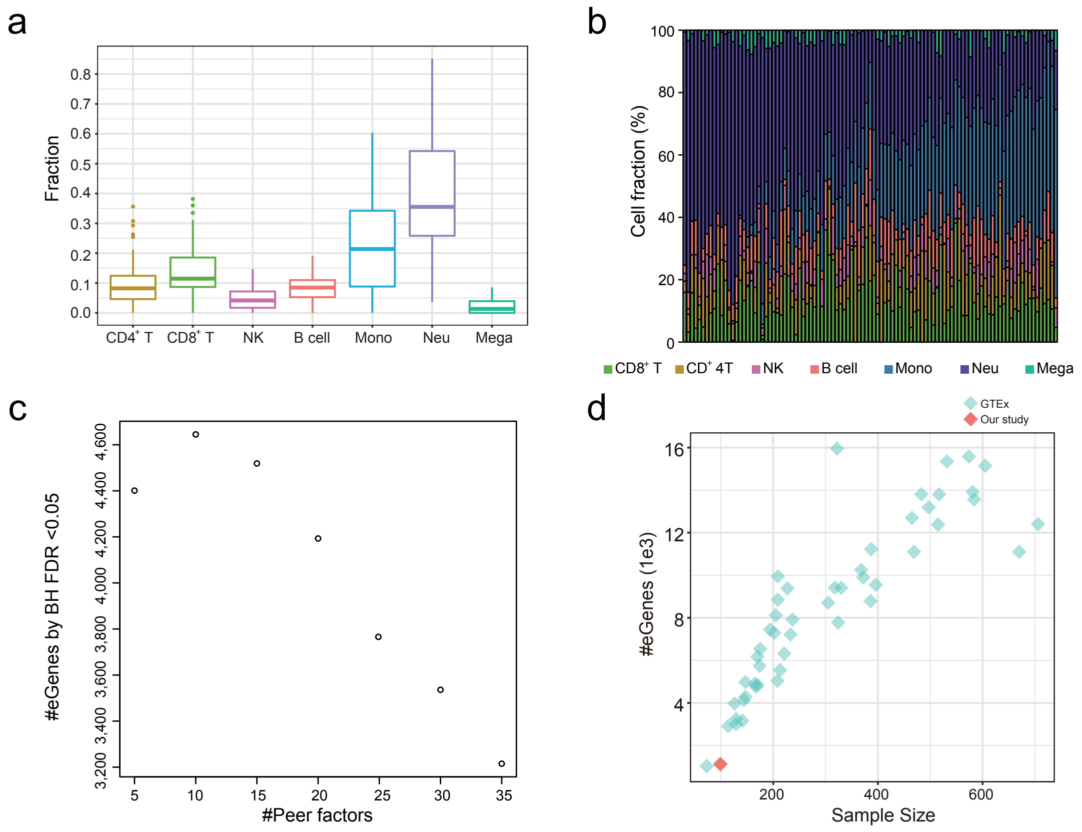


**Fig. S12. Deconvolution of immune cell fractions and eQTL optimization in LN whole blood samples.** (a) Boxplots showing estimated cell fractions of immune cell types in 99 LN whole blood samples using CIBERSORTx[55], with downsampled PBMC scRNA-seq data from 10 healthy controls and 11 LN patients. CD4T: CD4$^+$ T cells, CD8T: CD8$^+$ T cells, NK: natural killer cells, Mono: monocytes, Neu: neutrophils, Mega: megakaryocytes. (b) Distribution of relative cell fractions across 99 LN blood samples. (c) PEER factor optimization. The number of significant eQTLs (eGenes) depends on the number of included PEER factors. The eGenes were identified at a Benjamini-Hocheberg FDR < 0.05 to ensure the rapid optimization. (d) Relationship between sample size (x-axis) and the number of identified eGenes (y-axis). Turquoise diamonds represent the number of eGenes identified in 49 human tissues from GTEx (v8) [56], and the red diamond shows the number of eGenes identified in our LN whole blood eQTL dataset.

### Supplementary Figure 13

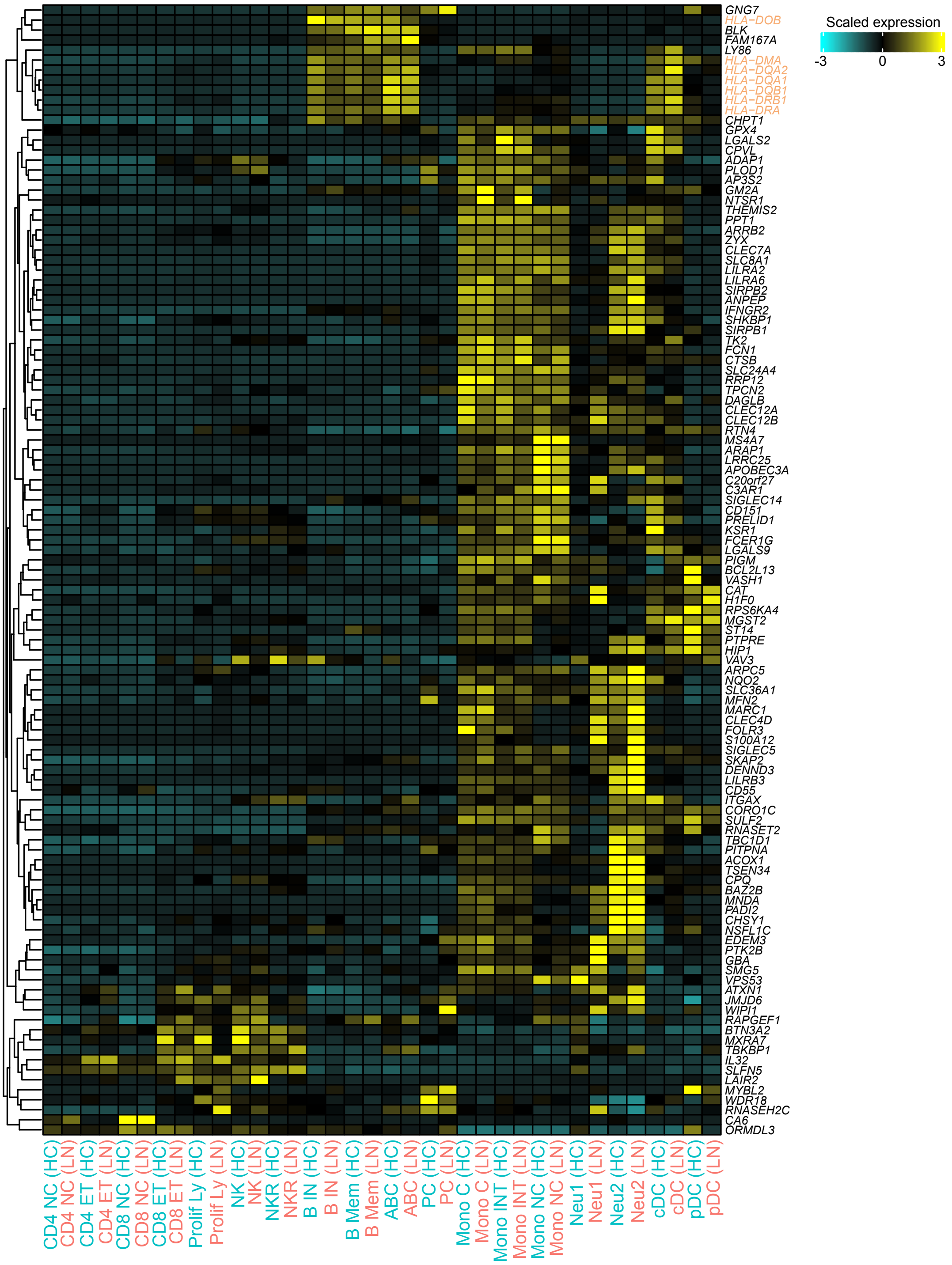


**Fig. S13. Expression patterns of 113 shared eGenes and HRGs across immune cell types in healthy controls (HCs) and LN patients.** Genes within the MHC region are highlighted in orange. Heatmap showing row-scaled expression of the 21 pro-inflammatory genes across 18 cell clusters (megakaryocytes excluded). The color scheme is based on Z-scores, calculated from normalized gene expression levels of the 18 cell types. Z-scores show the relatively expression levels compared to other clusters. Genes are hierarchically clustered based on expression profiles. CD4 NC: naïve CD4$^+$ T cells, CD4 ET: effector CD4$^+$ T cells, CD8 NC: naïve CD8$^+$ T cells, CD8 ET: effector CD8$^+$ T cells, Prolif Ly: proliferating lymphocytes, NK: natural killer cells, NKR: natural killer recruiting cells, B IN: naïve B cells, B Mem: memory B cells, ABC: age associated B cells, PC: plasma cells, Mono C: classical (CD14$^{++}$CD16$^-$) monocytes, Mono INT: intermediate (CD14$^{++}$CD16$^+$) monocytes, Mono NC: non-classical (CD14$^{dim}$CD16$^+$) monocytes, Neu1: neutrophil subpopulation 1, Neu2: neutrophil subpopulation 2, cDC: conventional dendritic cells, pDC: plasmacytoid dendritic cells.

**Supplementary Figure 14**

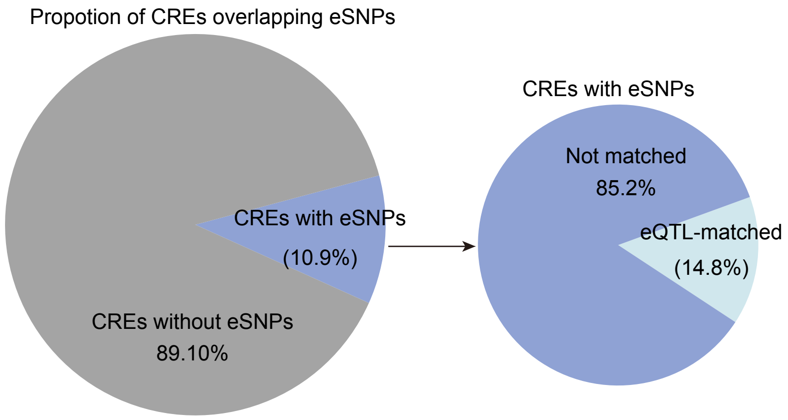


**Fig. S14. Overlap between peak-to-gene linkages and eQTLs.** *Left*: Pie chart showing the proportion of CREs overlapping at least one eQTL(cf) SNP. Right: Pie chart showing the proportion of these CREs linked to the same genes as the eGenes regulated by the overlapping SNPs.

### Supplementary Figure 15

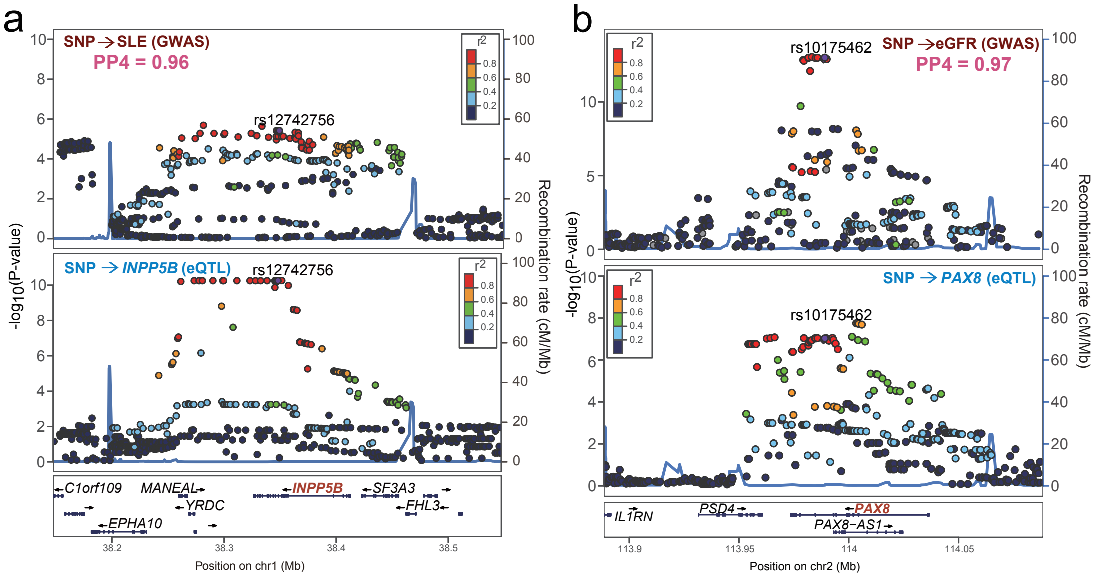


**Fig. S15. Multi-omic integrative annotation highlights the causal roles of *INPP5B* and *PAX8* in lupus nephritis.** (a) LocusZoom plots of SLE GWAS variants and blood *INPP5B* eQTL(cf)s. The *x* axis shows the ±100 kb genomic region around rs12742756. n = 12,653 individuals for SLE GWAS; n = 99 individuals for LN blood eQTL(cf)s. (b) LocusZoom plots of eGFR GWAS variants and blood *PAX8* eQTL(cf)s (*bottom*). n = 297,355 individuals for eGFR GWAS; n = 99 individuals for LN blood eQTL(cf)s. The *x* axis shows the ±100 kb genomic region around rs1017546. The *y* axis represents the significance -$\log_{10}$(two-sided P-value) of association tests (by linear regression). Each data point represents a variant, with color denoting the $r^2$ (the degree of LD).

### Supplementary Figure 16

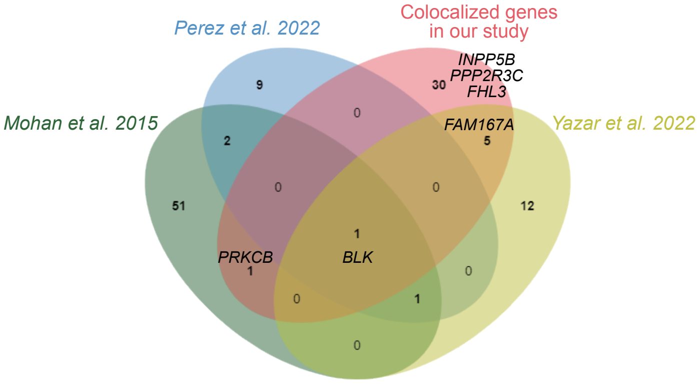


**Fig. S16. Venn diagrams showing the overlap of candidate causal genes for SLE and previously reported SLE/LN associated genes**[5, 57, 58]**.** Each circle represents the set of overlapped genes with SLE/LN associated genes reported in a specific study[5, 57, 58].

### Supplementary Figure 17

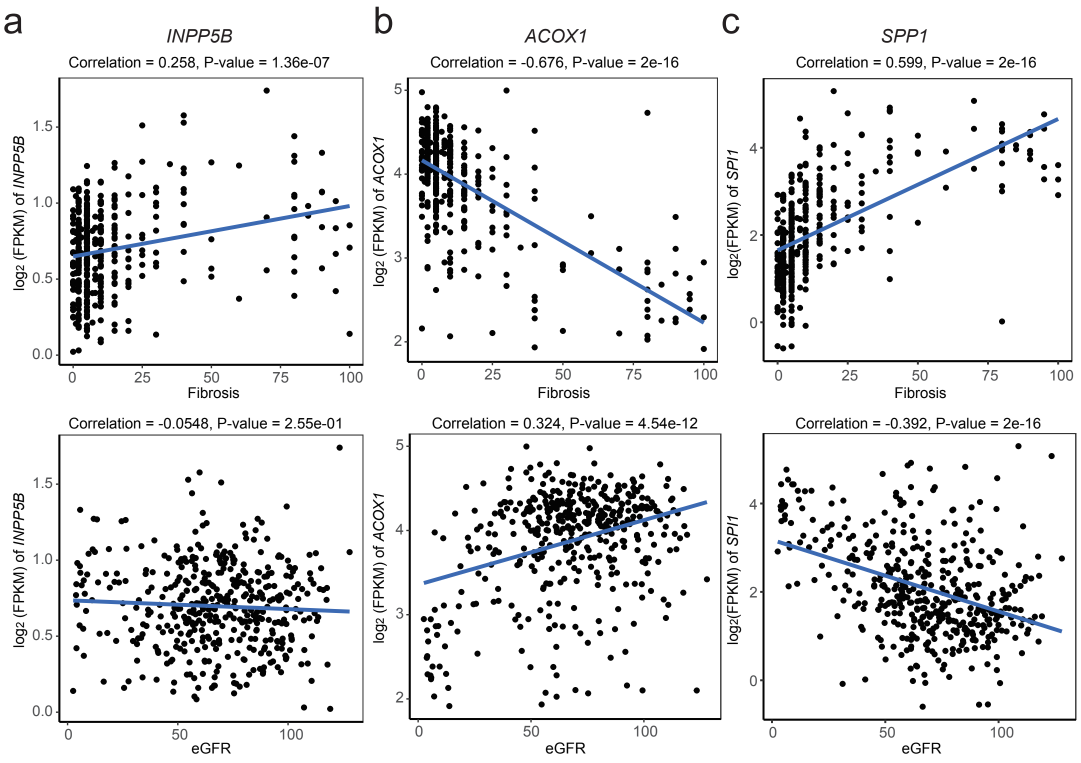


**Fig. S17. Correlation of selected gene expression with renal fibrosis and kidney function.** (a) Pearson's correlation of *INPP5B* expression with fibrosis (*top*) and eGFR (*bottom*) across 433 micro-dissected human kidney tubular samples. Two-sided P-value was calculated by t-test. (b) same as (a), but for *ACOX1*. (c) same as (a), but for *SPP1*.

### Supplementary Figure 18

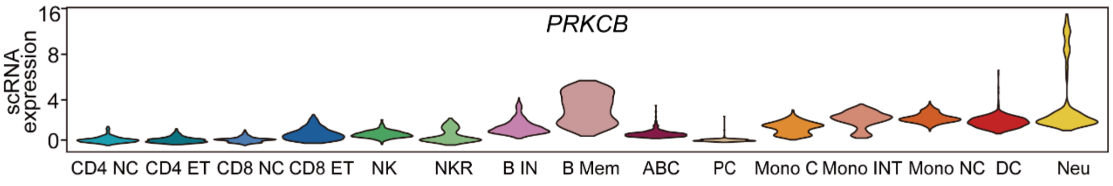


**Fig. S18. Violin plots showing *PRKCB* expression across immune cell types.** *Y*-axis shows log-normalized RNA expression values imputed by the *GeneIntegrationMatrix* function in ArchR from the snATAC-seq dataset. The width of each violin represents the density of cells at a given predicted expression level.

**Supplementary Figure 19**

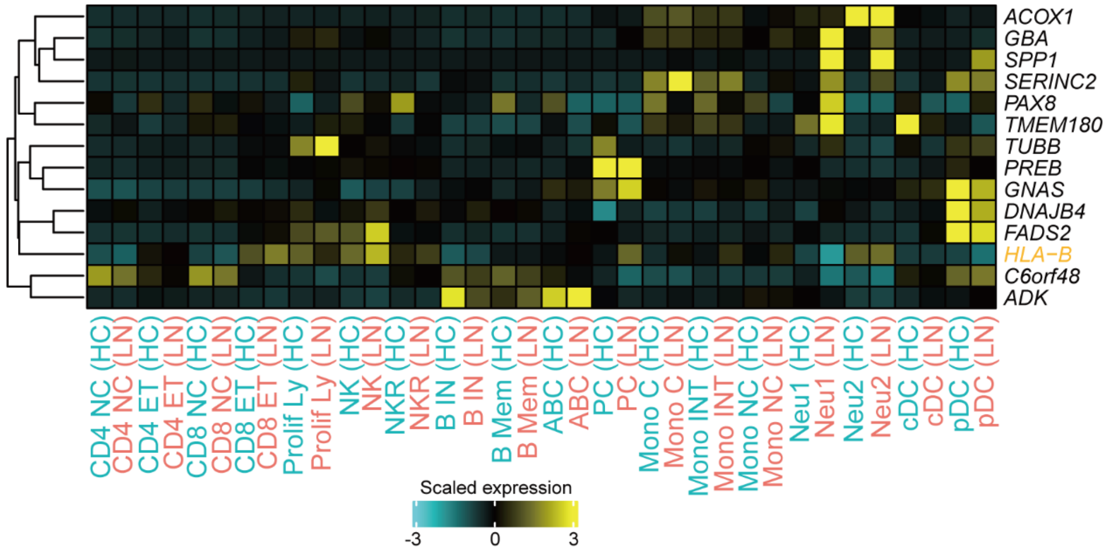


**Fig. S19. Expression patterns of 14 potential causal genes for eGFR across immune cell types in healthy controls (HCs) and LN patients.** Genes within the MHC region are highlighted in orange. Heatmap showing row-scaled expression of the 14 potential causal genes across 18 cell clusters (megakaryocytes excluded). The color scheme is based on Z-scores, calculated from normalized gene expression levels of the 18 cell types. Z-scores show the relatively expression levels compared to other clusters. Genes are hierarchically clustered based on expression profiles. CD4 NC: naïve $CD4^+$ T cells, CD4 ET: effector $CD4^+$ T cells, CD8 NC: naïve $CD8^+$ T cells, CD8 ET: effector $CD8^+$ T cells, Prolif Ly: proliferating lymphocytes, NK: natural killer cells, NKR: natural killer recruiting cells, B IN: naïve B cells, B Mem: memory B cells, ABC: age associated B cells, PC: plasma cells, Mono C: classical ($CD14^{++}CD16^-$) monocytes, Mono INT: intermediate ($CD14^{++}CD16^+$) monocytes, Mono NC: non-classical ($CD14^{dim}CD16^+$) monocytes, Neu1: neutrophil subpopulation 1, Neu2: neutrophil subpopulation 2, cDC: conventional dendritic cells, pDC: plasmacytoid dendritic cells..

### Supplementary Figure 20

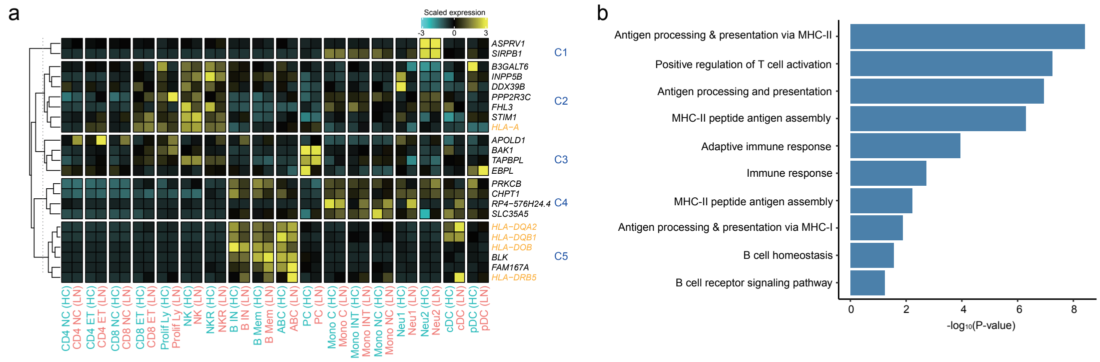


**Fig. S20. Expression patterns and functional annotation of 23 potential causal genes for SLE across immune cell types in healthy controls (HCs) and LN patients.** (a) Heatmap showing row-scaled expression of the 23 potential causal genes in PBMCs from LN patients across 18 cell clusters (megakaryocytes excluded). The color scheme is based on Z-scores, calculated from normalized gene expression levels of the 18 cell types. Genes within the MHC region are highlighted in orange. Z-scores show the relatively expression levels compared to other clusters. Genes are hierarchically clustered based on expression profiles (C1-C5). (b) Functional annotation (gene ontology, GO) of the 23 potential causal genes for SLE.

**Supplementary Figure 21**

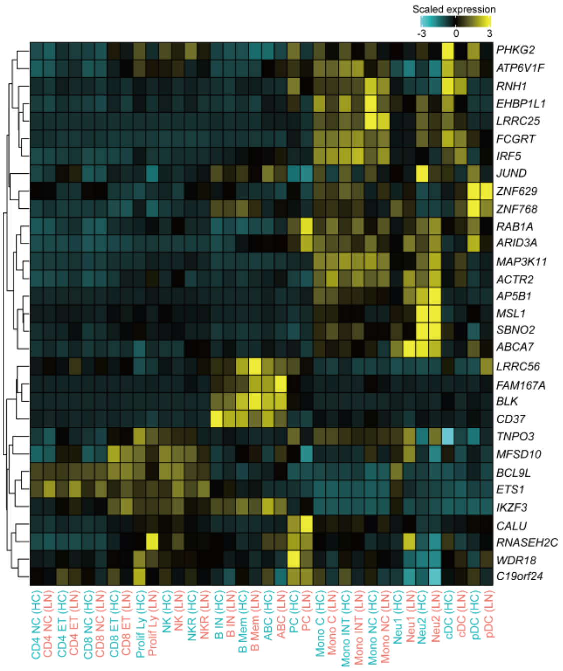


**Fig. S21. Expression patterns of 31 fmGWAS-linked genes shared between SLE and eGFR across immune cell types in healthy controls (HCs) and LN patients.** Heatmap showing row-scaled expression of the 31 fmGWAS-linked genes across 18 cell clusters (megakaryocytes excluded). The color scheme is based on Z-scores, calculated from normalized gene expression levels of the 18 cell types. Z-scores show the relatively expression levels compared to other clusters. Genes are hierarchically clustered based on expression profiles. CD4 NC: naïve $CD4^+$ T cells, CD4 ET: effector $CD4^+$ T cells, CD8 NC: naïve $CD8^+$ T cells, CD8 ET: effector $CD8^+$ T cells, Prolif Ly: proliferating lymphocytes, NK: natural killer cells, NKR: natural killer recruiting cells, B IN: naïve B cells, B Mem: memory B cells, ABC: age associated B cells, PC: plasma cells, Mono C: classical ($CD14^{++}CD16^-$) monocytes, Mono INT: intermediate ($CD14^{++}CD16^+$) monocytes, Mono NC: non-classical ($CD14^{dim}CD16^+$) monocytes, Neu1: neutrophil subpopulation 1, Neu2: neutrophil subpopulation 2, cDC: conventional dendritic cells, pDC: plasmacytoid dendritic cells.

### Supplementary Figure 22

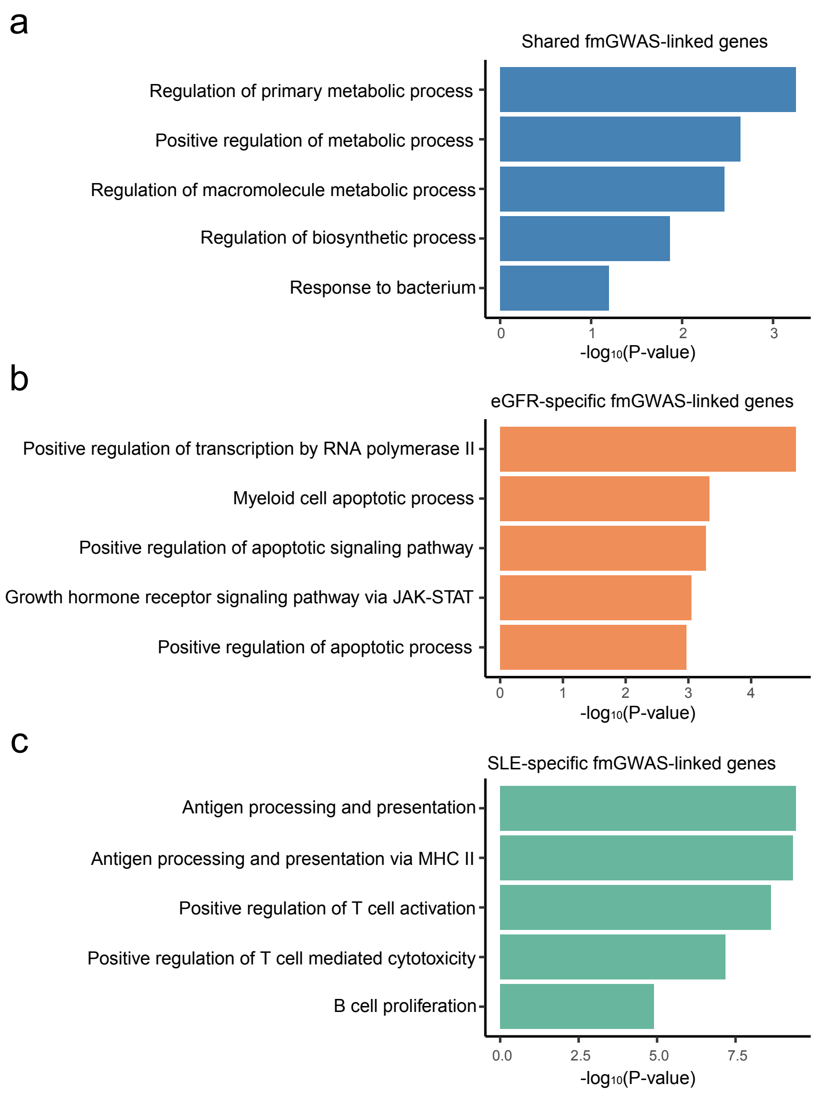


**Fig. S22. Functional annotation (gene ontology) of genes associated with fine-mapped variants for LN phenotypic manifestations, including SLE and eGFR GWAS.** Shown are enrichment results for (a) 31 genes shared between SLE and eGFR, (b) 380 eGFR fmGWAS-linked genes, and (c) 203 SLE fmGWAS-linked genes.

### Supplementary Figure 23

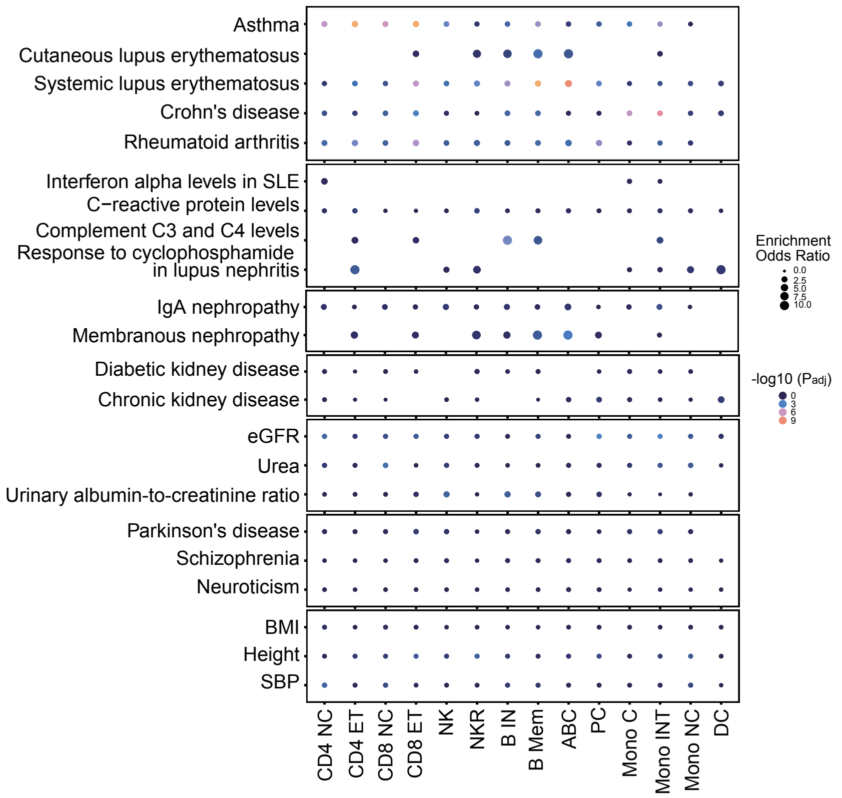


**Fig. S23. Enrichment of fine-mapped GWAS variants in cell type specific accessible chromatin regions.** Bubble plots show the enrichment of fine-mapped SNPs from SLE, eGFR, and other trait GWAS in cell type specific accessible regions. Dot size represents the odds ratio (OR), and dot color indicates statistical significance -$\log_{10}$(FDR-adjusted P-value). Enrichment was evaluated using a one-sided Fisher's exact test, comparing the overlap between fine-mapped SNPs for each trait (95% credible sets) with cell-type-specific accessible peaks relative to background peaks across all cell types.

**Supplementary Figure 24**

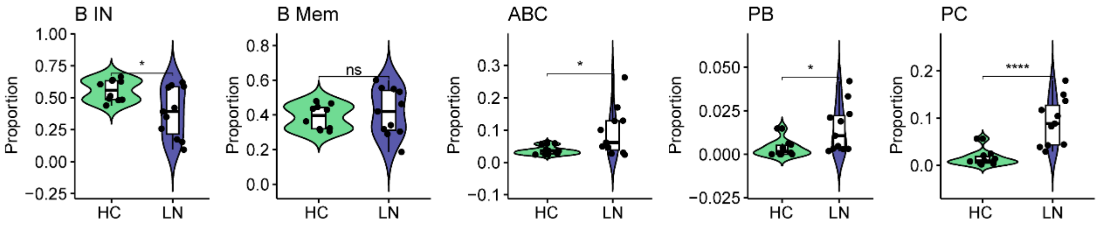


**Fig. S24. Violin plots showing fractions of B cell subsets in healthy controls (HCs, n = 10) and LN patients (n = 11).** P-values were calculated using the Wilcoxon test comparing group means between LN and HC. Significance levels are indicated as *p < 0.05, **p < 0.01, ***p < 0.001, and ****p < 0.0001. B IN: naïve B cells, B Mem: memory B cells, ABC: age associated B cells, PB: plasmablasts, PC: plasma cells.

### Supplementary Figure 25

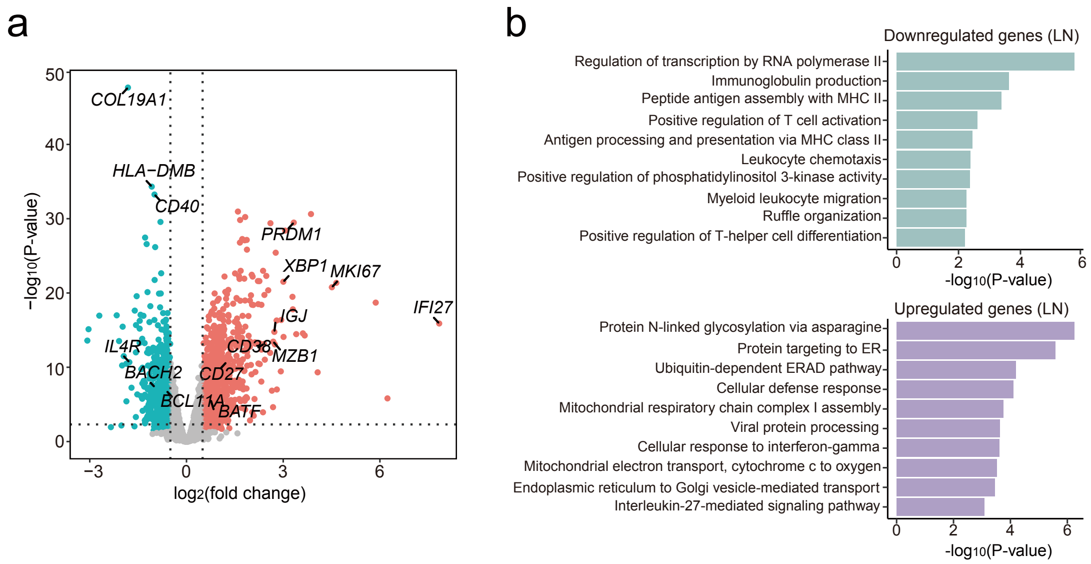


**Fig. S25. Differentially expressed genes in B cells between healthy controls and LN patients with functional annotation.** (a) Volcano plot of differentially expressed genes in B cells comparing LN patients (n = 11) and healthy controls (n = 10). *X*-axis represents the base 2 log of the fold change for gene expression (read counts) between B cells of LN patients and healthy controls. *Y*-axis is the negative based 10 of the association P-value. A total of 947 upregulated genes in LN are shown in red color, and 800 downregulated genes are shown in light blue color. The horizontal dashed line marks -$\log_{10}$(P-value) = 1, and vertical dashed lines mark |$\log_2$ fold change| = 0.5. Each data point represents an individual gene. (b) Functional annotation (gene ontology) of the top 500 downregulated (*top*) and upregulated (*bottom*) genes in B cells of 11 LN patients.

### Supplementary Figure 26

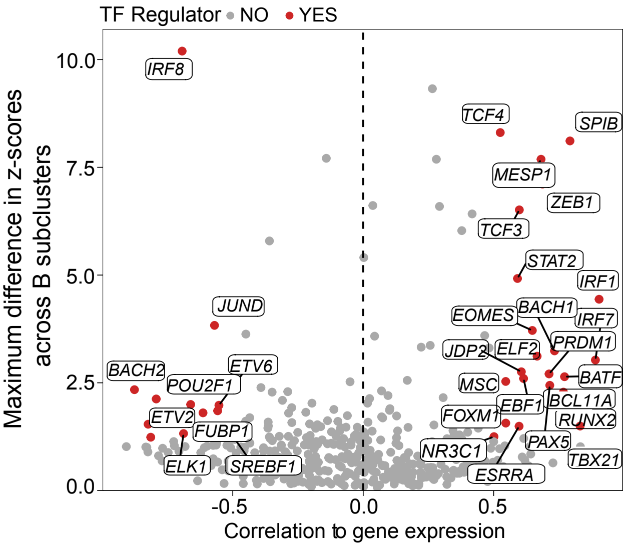


**Fig. S26. Correlation between TF motif activity and TF gene expression across B cell subclusters.** Scatter plot showing the correlation between TF motif activity, inferred from chromVAR[19] deviation z-scores, and corresponding TF gene expression across B cell subclusters. The y-axis represents the maximum difference in chromVAR deviation z-score between clusters, reflecting variability in motif activity, while the x-axis shows the Pearson's correlation coefficient between TF motif activity and TF gene expression across B cell subclusters. TFs with strong correlation (|Pearson's correlation| > 0.5) and high variability (top 25% of chromVAR deviation z-score differences) are highlighted in red.

**Supplementary Figure 27**

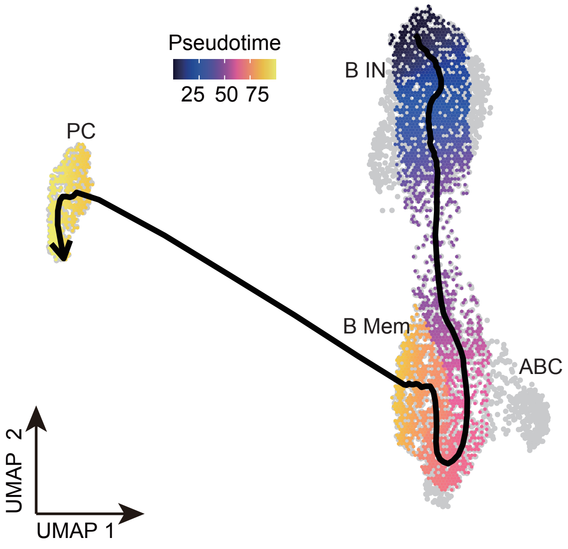


**Fig. S27. B cell trajectories and pseudotime analysis**. UMAP of B cells colored by pseudotime. The black line represents a double-spline-fitted trajectory along pseudotime. B IN: naïve B cells, B Mem: memory B cells, ABC: age associated B cells, PC: plasma cells.

### Supplementary Figure 28

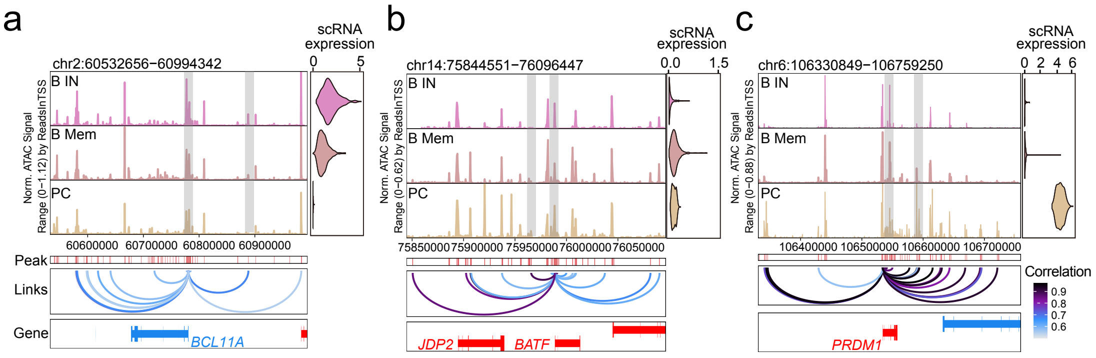


**Fig. S28. Chromatin accessibility and gene expression of representative TFs along the B cell differentiation trajectory.** Genomic tracks showing promoter regions of (a) *BCL11A*, (b) *BATF*, and (c) *PRDM1*. Right: violin plots of integrated gene expression across B cell types. Below (*top* to *bottom*): pseudobulk ATAC-seq peaks and peak-to-gene linkages (loop) identified from B cell subclusters. Gray vertical bars across both panels highlight selected open chromatin peaks exhibiting increased accessibility at specific stages of B cell differentiation.. B IN: Naïve B cells, B Mem: Memory B cells, PC: Plasma cells.

**Supplementary Figure 29**

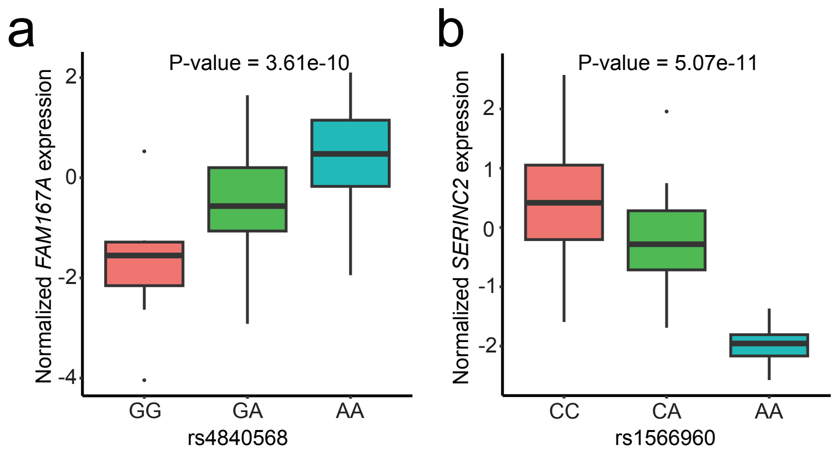


**Fig. S29. eQTL associations of SNPs with gene expression in LN whole blood samples.** (a) Association between the genotype of SNP rs4840568 and *FAM167A* gene expression in blood samples (n = 99). The *y* axis is the *FAM167A*-normalized expression in blood; the *x* axis is the genotype at the rs4840568 locus. (b) Association between the genotype of SNP rs1566960 and *SERINC2* gene expression in blood samples (n = 99). The *y* axis is the *SERINC2*-normalized expression in blood; the *x* axis is the genotype at the rs1566960 locus. The center lines show the medians; the box limits indicate the 25th and 75th percentiles; the whiskers extend to the 5th and 95th percentiles; the outliers are represented by the dots. The two-sided P-value was calculated using the linear regression eQTL(cf) model.

### Supplementary Figure 30

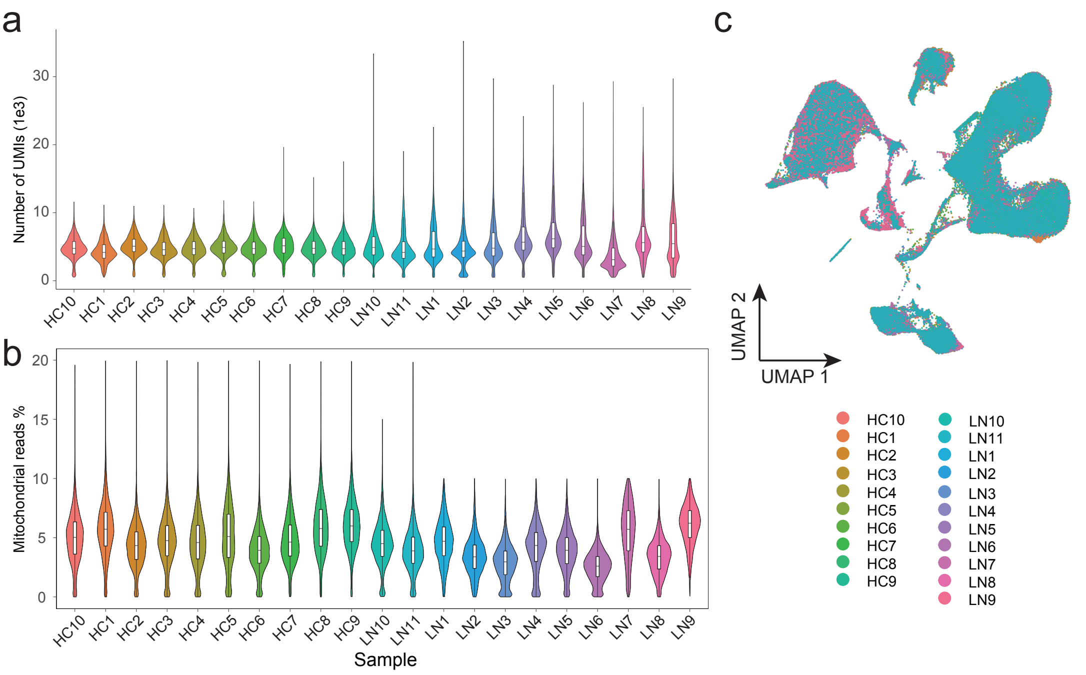


**Fig. S30. Quality control of human PBMC scRNA-seq data.** (a) Violin plots showing the number of unique reads per sample. The inset box plot indicates the median (center line), 25th and 75th percentiles (box), and whiskers extending to 1.5× the interquartile range. (b) Same as (a), but showing the percent of reads mapped to mitochondrial genes. (c) UMAP projection of the full scRNA-seq dataset after batch effect correction, colored by sample.

### Supplementary Figure 31

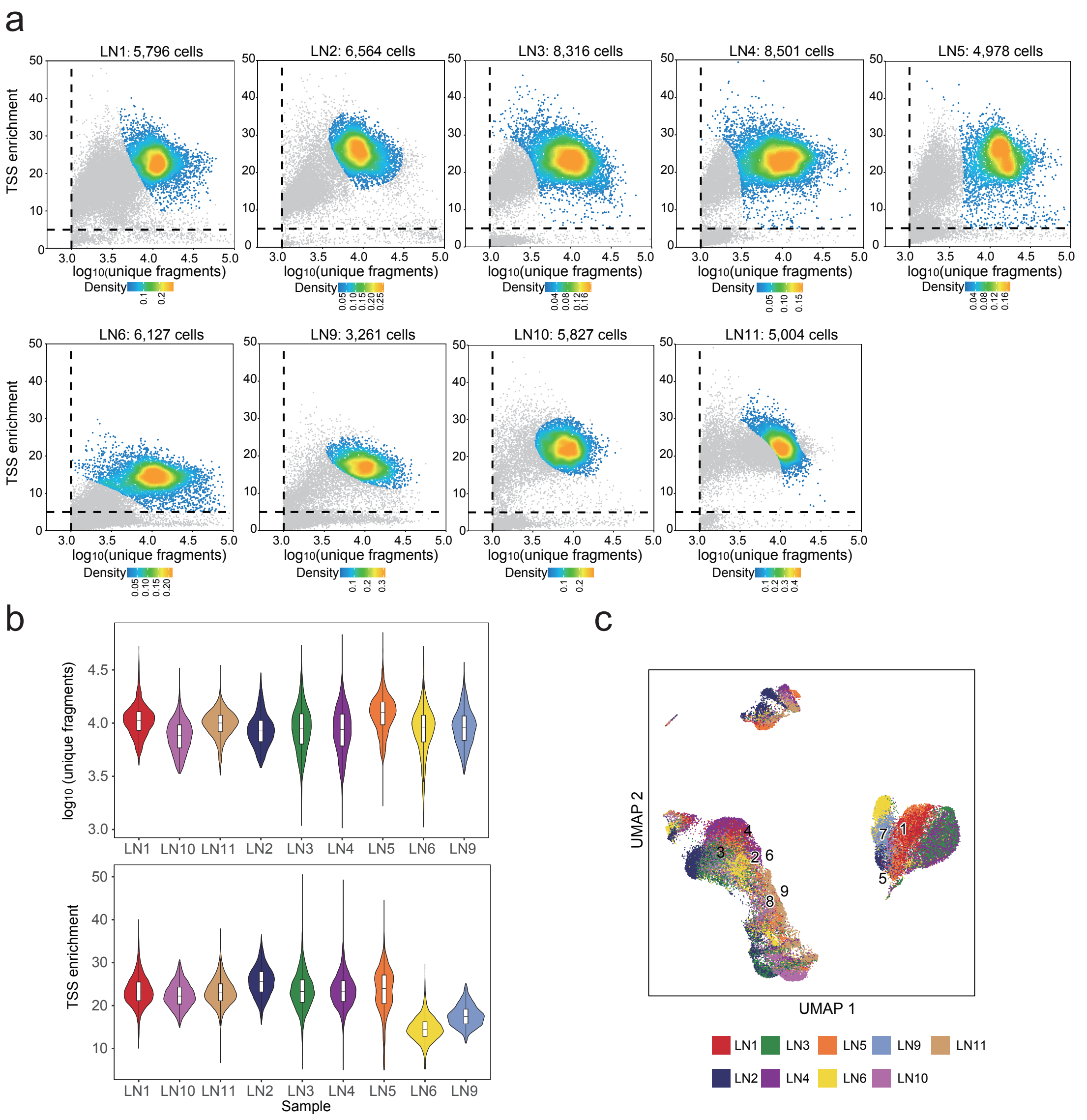


**Fig. S31. Quality control of human PBMC snATAC-seq data.** (a) Each data point represents a cell. The x-axis shows $\log_{10}$(unique fragments) per cell, and the y-axis shows the TSS enrichment score, reflecting read enrichment around known TSS regions. Gray dots indicate cells that did not pass quality-control filtering. (b) Violin plots showing the number of unique fragments (*top*) and TSS enrichment (*bottom*) for each snATAC-seq sample. Inset box plots indicate the median (center line), 25th and 75th percentiles (box), and whiskers extending to 1.5× the interquartile range. (c) UMAP projection of the full snATAC-seq dataset, colored by sample.
